# SBDH-Reader: an LLM-powered method for extracting social and behavioral determinants of health from clinical notes

**DOI:** 10.1101/2025.02.19.25322576

**Authors:** Zifan Gu, Lesi He, Awais Naeem, Pui Man Chan, Asim Mohamed, Hafsa Khalil, Yujia Guo, Jingwei Huang, Ismael Villanueva-Miranda, Ying Ding, Wenqi Shi, Matthew E. Dupre, Guanghua Xiao, Eric D. Peterson, Yang Xie, Ann Marie Navar, Donghan M. Yang

**Affiliations:** Quantitative Biomedical Research Center, Department of Health Data Science and Biostatistics, Peter O’Donnell Jr. School of Public Health, The University of Texas Southwestern Medical Center, Dallas, Texas, USA; Department of Health Data Science and Biostatistics, Peter O’Donnell Jr. School of Public Health, The University of Texas Southwestern Medical Center, Dallas, Texas, USA; School of Information, University of Texas at Austin, Austin, Texas, USA; The University of Texas Southwestern Medical Center, Dallas, Texas, USA; Department of Population Health Sciences, Duke University, Durham, North Carolina, USA; Department of Sociology, Duke University, Durham, North Carolina, USA; Department of Bioinformatics, The University of Texas Southwestern Medical Center, Dallas, Texas, USA; Simmons Comprehensive Cancer Center, The University of Texas Southwestern Medical Center, Dallas, Texas, USA; Department of Internal Medicine, The University of Texas Southwestern Medical Center, Dallas, TX, USA

## Abstract

**Objective:** Social and behavioral determinants of health (SBDH) are increasingly recognized as essential for prognostication and informing targeted interventions. Clinical notes often contain details about SBDH in unstructured format. Conventional extraction methods for these data tend to be labor intensive, inaccurate, and/or unscalable. In this study, we aim to develop and validate an LLM-powered method to extract structured SBDH data from clinical notes through prompt engineering.

**Materials and Methods:** We developed SBDH-Reader to extract six categories of granular SBDH data by prompting GPT-4o, including employment, housing, marital status, and substance use including alcohol, tobacco, and drug use. SBDH-Reader was developed using 7,225 notes from 6,382 patients in the MIMIC-III database (2001–2012) and externally validated using 971 notes from 437 patients at The University of Texas Southwestern Medical Center (UTSW; 2022–2023). We evaluated SBDH-Reader’s performance against human-annotated ground truths based on precision, recall, F1, and confusion matrix.

**Results:** When tested on the UTSW validation set, SBDH-Reader achieved a macro-average F1 ranging from 0.94 to 0.98 across six SBDH categories. For clinically relevant adverse attributes, F1 ranged from 0.96 (employment; housing) to 0.99 (tobacco use). When extracting any adverse attributes across all SBDH categories, SBDH-Reader achieved an F1 of 0.97, recall of 0.97, and precision of 0.98 in the independent validation set.

**Conclusion:** A general-purpose LLM can accurately extract structured SBDH data through effective prompt engineering. The SBDH-Reader has the potential to serve as a scalable and effective method for collecting real-time, patient-level SBDH data to support clinical research and care.

## BACKGROUND AND SIGNIFICANCE

Social and behavioral determinants of health (SBDH) are increasingly recognized as essential for understanding a patient’s clinical presentation and as targets for interventions.^1–4^ SBDH encompass a broad range of factors that describe an individual’s living conditions at the personal, community, and societal levels. A growing body of research highlights the significance of integrating SBDH to enhance clinical risk assessment, outcome prediction, and clinical trial enrollment, ultimately guiding therapeutic interventions across various clinical settings.^4–11^

Despite the recognized significance of SBDH in the medical domain, access to these patient-level characteristics is limited due to the lack of effective methods for high-quality and continuous data collection, particularly in clinical settings.^12,13^ Previous studies have attempted to collect these data through patient surveys delivered during routine care, but the scalability and generalizability of this approach is limited.^14,15^ Many electronic health records (EHRs) provide structured data fields for SBDH, but these require providers to enter and are often poorly populated.^12,13,16^ Information about SBDH is often documented in the narrative sections of clinical notes, but extracting detailed SBDH information from unstructured notes has proven challenging.^4,12,13,17–20^

Various natural language processing (NLP) methods have been applied to identify SBDH from clinical notes, ranging from rule-based approaches to deep learning-based models, including early language models such as BERT.^18,21–31^ However, conventional NLP approaches fall short in SBDH extraction tasks due to limitations in accuracy, requirement of entity/relation-level annotations, and scalability to real-world settings.^18,24,32,33^ Recently, large language models (LLMs) have demonstrated substantial potential in healthcare and biomedical applications,^34–39^ particularly for extracting information from clinical notes.^40–42^ Unlike traditional NLP approaches, LLMs excel in text extraction tasks without the need for task-specific model (pre)training or detailed, entity- and relation-level annotations.^18,32,34,43^ For example, the 2022 n2c2/UW NLP challenge on extracting social determinants of health produced an extensively annotated corpus containing 18,000 distinct events, each with multiple annotated arguments, resulting in a massive and valuable resource for developing traditional NLP methods.^24,32^ In contrast, recent LLM-based studies on SBDH often rely on simpler sentence- or note-level annotations, typically involving a few hundred to a few thousand records.^29–31^

LLMs operate by processing sequential text as input and predicting the next token based on their training. Prompting is the primary method of directing LLMs to generate desired outputs by providing detailed instructions in the form of a prompt, allowing the model to predict the appropriate sequence of tokens in response. Prompt engineering—the practice of crafting high-quality prompts that effectively guide LLMs to generate target outputs—is a crucial step in adapting general-purpose LLMs for specific tasks.^44^ This approach enables LLMs to perform a wide range of tasks, including text summarization, question answering, code generation, and language translation, without requiring model fine-tuning. For the specific task of extracting information from clinical notes, the prompt includes (a) comprehensive and precise instructions for the extraction task and (b) the raw clinical text that serves as the source for information extraction.

To date, the application of prompt engineering-based LLMs to SBDH data extraction remains largely unexplored. Existing studies are limited by their restriction to sentence-level input ^25,29–31^, lack of precise evaluation of individual SBDH categories^31,45^, and a classifier-only design that requires model training or fine-tuning towards predefined, fixed SBDH coding.^25,30,31,45^ Unlike medical concepts, which often fall under specialized domains, descriptions of SBDH in clinical notes typically require less domain knowledge and terminology, a foundation on which advanced LLMs are well-equipped. On the other hand, categorization and coding of SBDH can vary significantly across health systems, departments, and author types. Therefore, the conventional strategy of training or fine-tuning models to fit predefined SBDH targets is challenged. Prompt engineering-based strategies may offer a more flexible, versatile, and generalizable solution.

## OBJECTIVE

In this study, we aim to develop SBDH-Reader, an LLM-powered, prompting-based method for extracting SBDH data from real-world clinical notes. We specifically explore whether advanced LLMs, such as GPT-4o, can achieve high accuracy in identifying SBDH without fine-tuning. Through prompting, we task the SBDH-Reader with determining the granular attribute of a given SBDH category by processing full-length SBDH documentation. For example, for the SBDH category of “employment”, SBDH-Reader extracts all supporting information from a note to determine the appropriate attribute among “employed”, “unemployed”, “underemployed”, “retired”, and “student”. Overall, the SBDH-Reader performs multi-label (i.e., across multiple SBDH categories) and multi-class classification (i.e., across multiple possible attributes) in a single prompt, highlighting its potential as a scalable and effective tool for collecting real-world SBDH data to support clinical research and care.

## MATERIALS AND METHODS

### Datasets and Data Processing

This study involves three EHR-based datasets with clinical notes (Table 1). For method development, two previously established datasets based on the Medical Information Mart for Intensive Care III (MIMIC-III) database^46^ were used, which contains data from 2001 to 2012. The first dataset, established by Guevara et al. (MIMIC-G)^29^, contains 200 notes written by physicians, nurses, and social workers for 183 patients. To generate full-length notes as input data for SBDH-Reader, we merged the original sentence-level texts (5328 entries) and ground truths from the MIMIC-G dataset into the note level (200 entries). The second dataset, established by Ahsan et al. (MIMIC-A)^47^, contains 7025 discharge summaries for 6199 patients. Following the original annotation method, we only used “social history” section as input data.^47^

**Table 1.** Cohort characteristics. P-value was based on chi-square test for patient-level characteristics between each MIMIC-III dataset and the UTSW dataset.

|  | MIMIC-G | MIMIC-A | UTSW |
| --- | --- | --- | --- |
| Role | Development | Development | Testing |
| **Patient-level Characteristics** |  |  |  |
| Number of patients | 183 | 6199 | 437 |
| Sex | P < 0.05 | P < 0.05 |  |
| Male | 101 (55.2%) | 2687 (43.3%) | 212 (48.5%) |
| Female | 82 (44.8%) | 3512 (56.7%) | 225 (51.5%) |
| Race | P < 0.05 | P < 0.05 |  |
| White | 131 (71.6%) | 4496 (72.5%) | 241 (55.1%) |
| Black | 16 (8.7%) | 540 (8.7%) | 144 (33.0%) |
| Other | 9 (4.9%) | 301 (4.9%) | 10 (2.3%) |
| Unknown | 27 (14.8%) | 862 (13.9%) | 42 (9.6%) |
| Ethnicity | P < 0.05 | P < 0.05 |  |
| Non-Hispanic | 156 (85.2%) | 5337 (86.1%) | 348 (79.6%) |
| Hispanic | 12 (6.6%) | 214 (3.4%) | 61 (14.0%) |
| Unknown | 15 (8.2%) | 648 (10.5%) | 28 (6.4%) |
| **Note-level Characteristics** |  |  |  |
| Number of notes | 200 | 7025 | 971 |
| Number of notes by type (n) | Nursing (65)<br>Physician (70)<br>Social work (65) | Discharge summary (7025) | Consults (372)<br>H&P (333)<br>Progress (230)<br>ED Provider (36) |
| **Number of notes with SBDH information** |  |  |  |
| Employment | 36 (18.0%) | 2730 (38.9%) | 395 (40.7%) |
| Housing | 2 (1.0%) | 4429 (63.0%) | 259 (26.7%) |
| Marital relationship | 63 (31.5%) | N.A. | 843 (86.8%) |
| Alcohol use | N.A. | 5036 (71.7%) | 909 (93.6%) |
| Tobacco use | N.A. | 5379 (76.6%) | 921 (94.9%) |
| Drug use | N.A. | 2564 (36.5%) | 897 (92.4%) |

N.A.: The ground truths for the SBDH category was not provided in the published dataset.

The independent validation dataset was derived from the EHRs at The University of Texas Southwestern Medical Center (UTSW). We established this dataset from a heart failure cohort of 437 patients treated at UTSW, containing 971 full-length notes of various types dated from 2022 to 2023, including progress, history and physical (H&P), consult, emergency department (ED) provider notes. To meaningfully evaluate SBDH-Reader’s performance and mitigate the intrinsic data imbalance caused by missing SBDH documentation, we applied keyword-based filtering to all available notes for this cohort in the UTSW EHRs, yielding a dataset enriched with SBDH documentation (Supplementary Info 1). Similarly, we extracted the “social history” section as input data.

Data from MIMIC-III were de-identified at the source.^46^ Data from UTSW were de-identified in house. The LLM used in this study, GPT-4o, was operated on UTSW’s private Microsoft Azure OpenAI service. Per contractual agreements with Microsoft, data processed on this service will remain inaccessible to OpenAI or other customers and will not be used to improve OpenAI models or any Microsoft or 3rd party products or services. Use of MIMIC-III data in this setting is in compliance with the PhysioNet Credentialed Health Data Use Agreement 1.5.0^48^ and the policy of “Responsible use of MIMIC data with online services like GPT”^49^. This study protocol was approved by UTSW’s institutional review board (Protocol #STU-2024-0087).

### Task Definition and Data Annotation

The task of extracting SBDH from a given note was defined as a multi-label, multi-class classification problem, where each label corresponds to an SBDH category. This study targets six SBDH categories: employment, housing, marital relationship, and substance use including alcohol, tobacco, and drug use. An SBDH terminology was constructed by integrating the terminologies used in the two MIMIC-based studies ^29,47^ and the UTSW Epic system, in consultation with a medical sociologist (MED) and two physicians (AMN, EDP) (Table 2). This terminology defines the permissible attributes in each category. Notably, the terminologies defined in the MIMIC-G and MIMIC-A studies resulted in ground truth annotations with varying granularities and standards, as commonly seen in the field^25,29–31,45,47^. To ensure consistency across different standards, we tasked the SBDH-Reader with classifying only at the granular attribute level defined in Table 2, without allowing the LLM to determine these as positive or negative attributes. Then, to enable evaluation against the original attributes used in the MIMIC-G and MIMIC-A studies^29,47^, we applied a rule-based mapping of the SBDH-Reader-identified attributes to three post hoc classes—adverse attributes, non-adverse attributes, or no information (Table 2).

**Table 2.**
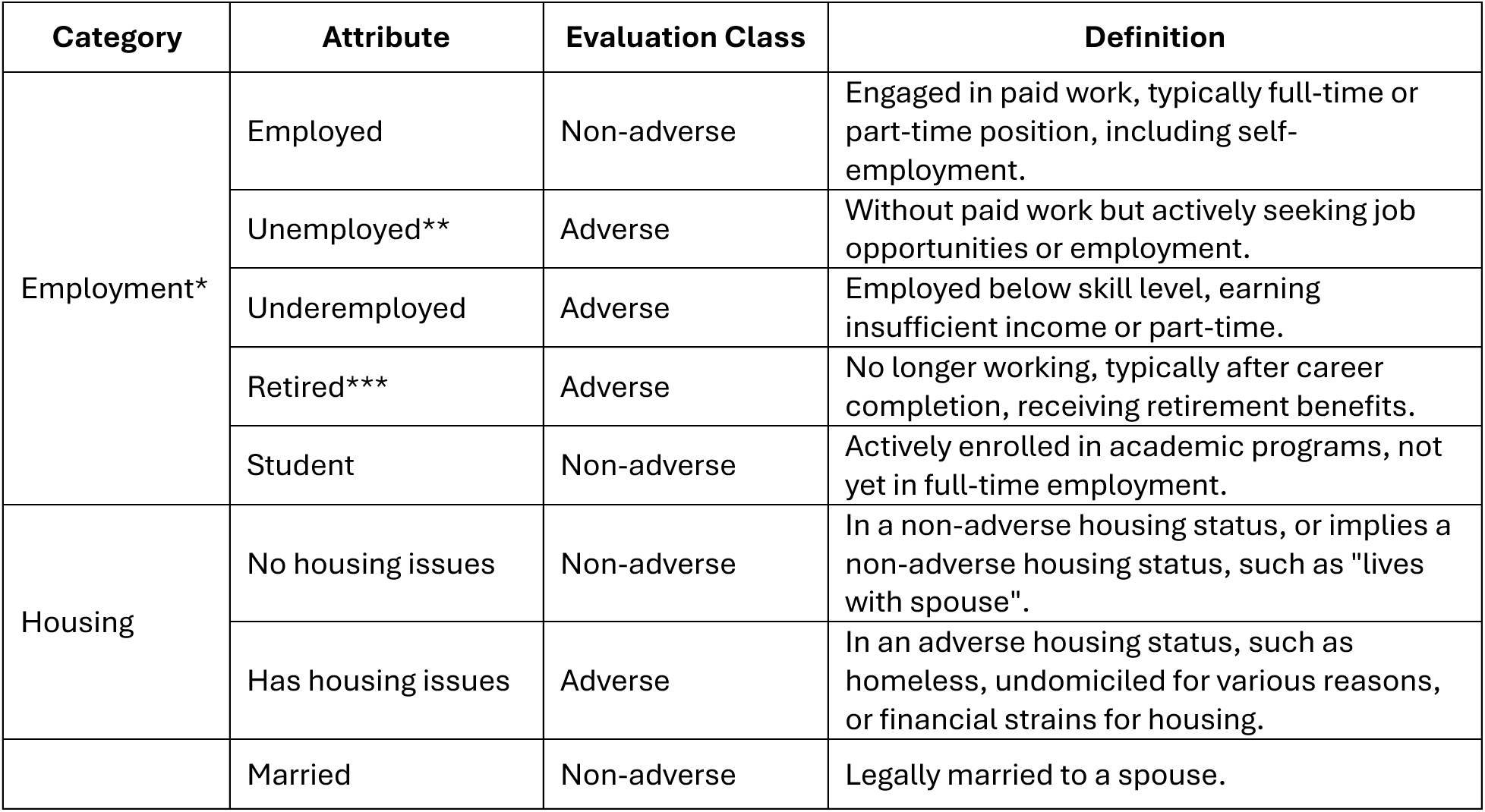
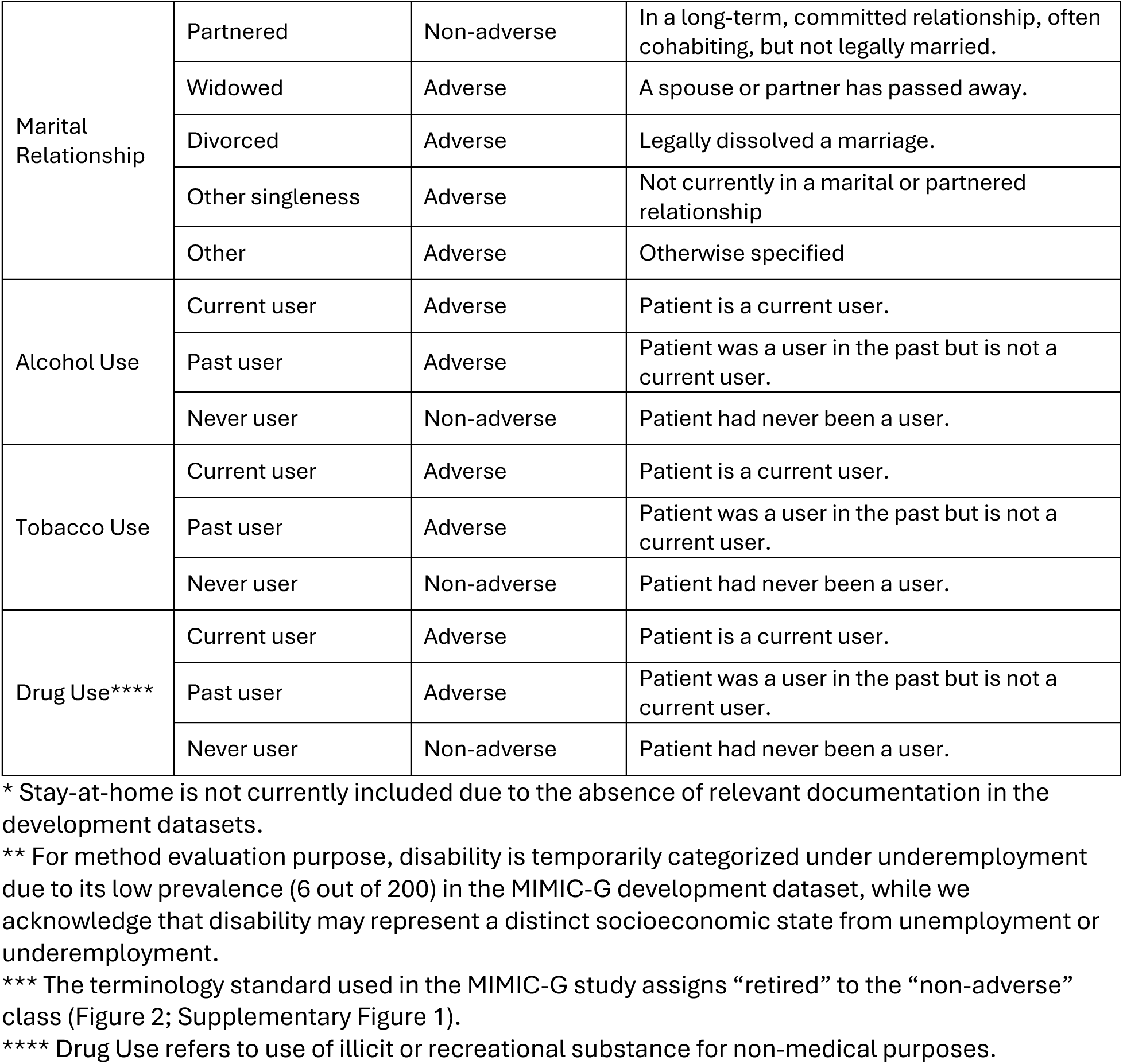
SBDH terminology. Each category also allows a universal “no information” attribute when no explicit evidence of other attributes is identified. SBDH-Reader is designed to predict granular attributes rather than aggregated classes (i.e., adverse or non-adverse). The mapping of attributes to classes is applied solely for meaningful quantitative evaluation, given the relatively small sample size for many individual attributes and the predominance of the “no information” attribute. This assignment is not intended to reflect a universal or absolute categorization, as the interpretation of each attribute may vary depending on specific clinical or research contexts. The two-level design of SBDH-Reader—attribute-level prediction and class-level evaluation—preserves the model’s capacity to identify precise SBDH features while enabling adjustable aggregation of such features that better support downstream analysis.

All human annotations in this study followed the same two-step approach: first, annotating the granular attributes, and then applying rule-based mapping to the three evaluative classes. To generate ground truths for the UTSW datasets, two medical students (AM, HK) annotated all notes under the supervision of two data scientists (LH, DMY) and one physician (AMN) using a local instance of the INCEpTION platform^50^ (see detailed annotation guidelines in Supplementary Info 2). Initially, the two annotators practiced on the first 100 notes in the UTSW datasets, discussed the results with the supervisors, and repeated the process. Once a good agreement was achieved (Cohen’s Kappa ≥ 0.85, Supplementary Table 1), they proceeded with independent annotation of the entire dataset. Any remaining discrepancies were resolved by LH and DMY to generate the final ground truths.

To ensure consistent annotation standards across all three datasets, after the annotation of the UTSW datasets, three data scientists (ZG, LH, DMY) conducted a thorough review of the original annotations provided with the two MIMIC datasets following the in-house established annotation guidelines. Mislabelings were corrected to generate the final ground truths for the MIMIC-G and MIMIC-A used in this study. This ground truth review and correction process was conducted in parallel with the iterative prompt refinement process (see the next section).

### Model and Prompt Engineering

We used GPT-4o (version 2024-05-13) in this study. Through prompting, the LLM was instructed to process an entire note entry and summarize and classify all relevant SBDH descriptions into pre-defined categories and attributes based on the established terminology (Figure 1). Each note was processed in a single-round chat completion, with all involved SBDH categories handled in one prompting.

**Figure 1.**
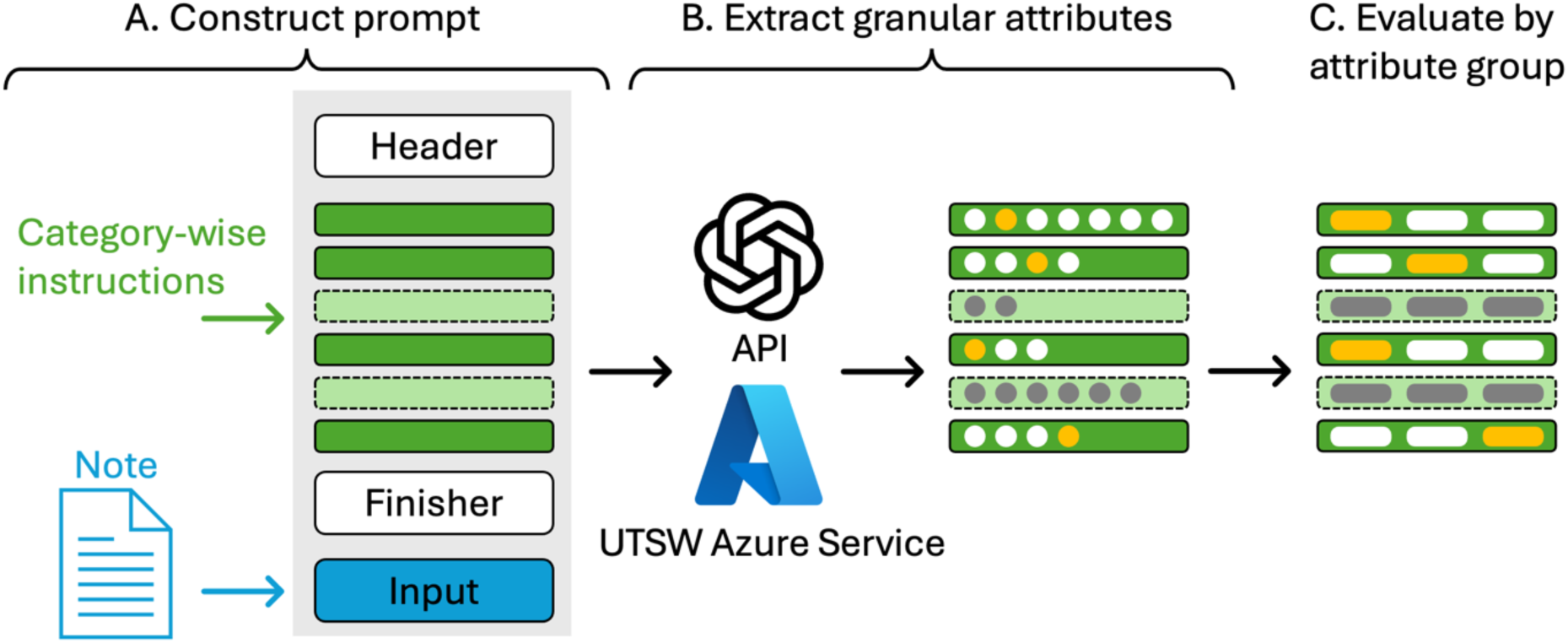
Overview of SBDH-Reader. The three key components of developing and validating SBDH-Reader are: (A) prompt construction, (B) result generation, and (C) evaluation. (A) The LLM prompt for SBDH-Reader is structured into a global header, category-wise instructions (green boxes), a global finisher, and input note (blue box). Each dark green box with a solid edge represents instructions for an included SBDH category, while each light green box with a dashed-line edge represents instructions available in SBDH-Reader but skipped during a given LLM query based on specific operational conditions (e.g., data, task, etc.). (B) The constructed prompt is sent to the UTSW private Azure OpenAI service via API to classify each included SBDH category into granular attributes (yellow circles). All circles represent permissible granular attributes for the LLM to target, while gray circles indicate skipped generations due to excluded SBDH categories (light green boxes). (C) For evaluation, granular attributes are mapped onto three attribute groups— adverse attributes, non-adverse attributes, and no information—using a rule-based mapping (Supplementary Table 1).

The SBDH-Reader prompt consists of two sections: instruction and input (Figure 1; Supplementary Info 3). The input section corresponds to a single note entry. The instruction section follows a modular design by sequentially listing each involved SBDH category along with category-specific definitions of permissible attributes and detailed descriptions. For example, the instruction under “employment” defines six permissible attributes: employed, unemployed, underemployed, retired, student, and no information (Table 2). Definition and specification surrounding each attribute is provided along with the prompt. For instance, under Alcohol Use we specified that any mention of polysubstance abuse implies alcohol use unless otherwise stated. To ensure structured outputs, we prompted the LLM to generate responses in JSON format, where the first key represents the identified attribute for each category, and the second key provides supporting evidence extracted from the text. The JSON format was further enforced by specifying the *response_format* parameter within the Azure OpenAI API. Last, a global finisher defines general instructions applicable across all categories. For example, we explicitly instructed the LLM to prioritize the current SBDH status at the time of the note, and not focus on any speculative statements about future changes (Supplementary Info 3).

We developed and iteratively refined the prompt using the MIMIC-G and MIMIC-A datasets. This process involved thorough error analysis, during which we added prompt specifications at both the category and attribute levels to address observed errors. The refinement process was concluded when minimal improvement was observed between iterations. During this same process, we reviewed and corrected inaccuracies in the original ground truths provided in the MIMIC-G and MIMIC-A datasets. Various milestone versions of the prompt developed during this process are presented in Supplementary Info 4. Detailed procedures and results of this iterative prompt refinement and ground truth correction process are provided in Supplementary Info 5 and Supplementary Tables 2 and 3. In independent testing with the UTSW dataset, no prompt changes were allowed.

### Evaluation and Error Analysis

The metrices we used to evaluate SBDH-Reader’s performance include precision (positive predictive value), recall (sensitivity), F1, and confusion matrix. Considering the high imbalance across various attributes, including “no information”, in all three datasets, we present the evaluations on the level of three combined classes: adverse attributes, non-adverse attributes, or no information. For precision, recall, and F1, we first report macro-average across these three classes for each SBDH category to show general performance. Then, we report these metrics on the adverse attributes alone to show its performance in this clinically relevant class (Table 3). Median and 95% confidence interval were estimated by bootstrapping with 500 iterations, sampling the entire dataset with replacement. The housing category from MIMIC-G was not evaluated due to low SBDH documentation rate (99% missing, Table 1). Error analysis was conducted by characterizing key error patterns through a case-by-case review of all misclassifications made by SBDH-Reader on the UTSW dataset.

**Table 3.** Performance of SBDH-Reader on development and validation datasets. Note-level results are presented as median followed by (lower–upper) bounds of the 95% confidence interval, estimated by bootstrapping with 500 iterations, sampling the entire dataset with replacement.

|  | Macro average |  |  | Adverse attributes only |  |  |
| --- | --- | --- | --- | --- | --- | --- |
| Category | F1 | Precision | Recall | F1 | Precision | Recall |
| MIMIC-G |  |  |  |  |  |  |
| Employment | 0.98<br>(0.94–1.00) | 0.96<br>(0.91–1.00) | 0.99<br>(0.99–1.00) | 1.00<br>(1.00–1.00) | 1.00<br>(1.00–1.00) | 1.00<br>(1.00–1.00) |
| Marital Relationship | 0.90<br>(0.83–0.95) | 0.89<br>(0.82–0.95) | 0.92<br>(0.85–0.96) | 0.84<br>(0.69–0.95) | 0.75<br>(0.56–0.94) | 0.94<br>(0.79–1.00) |
| Overall* | 0.94<br>(0.90–0.96) | 0.93<br>(0.89–0.96) | 0.95<br>(0.92–0.98) | 0.92<br>(0.84–0.98) | 0.86<br>(0.78–0.97) | 0.97<br>(0.89–1.00) |
| MIMIC-A |  |  |  |  |  |  |
| Employment | 0.83<br>(0.83–0.84) | 0.82<br>(0.81–0.83) | 0.86<br>(0.85–0.87) | 0.84<br>(0.83–0.85) | 0.80<br>(0.78–0.82) | 0.88<br>(0.86–0.89) |
| Housing | 0.91<br>(0.89–0.92) | 0.90<br>(0.88–0.93) | 0.92<br>(0.90–0.93) | 0.88<br>(0.83–0.92) | 0.83<br>(0.76–0.89) | 0.93<br>(0.88–0.98) |
| Alcohol Use | 0.91<br>(0.91–0.92) | 0.93<br>(0.92–0.94) | 0.90<br>(0.90–0.91) | 0.93<br>(0.92–0.93) | 0.89<br>(0.87–0.90) | 0.98<br>(0.97–0.98) |
| Tobacco Use | 0.94<br>(0.93–0.94) | 0.95<br>(0.95–0.96) | 0.93<br>(0.92–0.93) | 0.97<br>(0.96–0.97) | 0.95<br>(0.94–0.95) | 0.99<br>(0.98–0.99) |
| Drug Use | 0.86<br>(0.85–0.87) | 0.83<br>(0.82–0.85) | 0.90<br>(0.89–0.91) | 0.79<br>(0.75–0.81) | 0.71<br>(0.67–0.76) | 0.87<br>(0.84–0.90) |
| Overall* | 0.90<br>(0.90–0.91) | 0.90<br>(0.89–0.91) | 0.91<br>(0.90–0.91) | 0.89<br>(0.88–0.90) | 0.86<br>(0.84–0.88) | 0.93<br>(0.92–0.94) |
| UTSW |  |  |  |  |  |  |
| Employment | 0.94<br>(0.93–0.96) | 0.97<br>(0.96–0.98) | 0.93<br>(0.90–0.95) | 0.96<br>(0.94–0.98) | 0.94<br>(0.92–0.97) | 0.98<br>(0.96–0.99) |
| Housing | 0.95<br>(0.93–0.98) | 0.95<br>(0.90–0.97) | 0.96<br>(0.93–0.98) | 0.96<br>(0.88–1.00) | 0.96<br>(0.83–1.00) | 0.96<br>(0.85–1.00) |
| Marital Relationship | 0.98<br>(0.96–0.99) | 0.97<br>(0.96–0.99) | 0.98<br>(0.97–0.99) | 0.98<br>(0.97–0.99) | 0.98<br>(0.96–0.99) | 0.99<br>(0.99–1.00) |
| Alcohol Use | 0.97<br>(0.96–0.99) | 0.96<br>(0.93–0.98) | 0.99<br>(0.98–0.99) | 0.98<br>(0.98–0.99) | 0.99<br>(0.99–1.00) | 0.97<br>(0.95–0.98) |
| Tobacco Use | 0.98<br>(0.97–1.00) | 0.97<br>(0.95–0.99) | 1.00<br>(0.99–1.00) | 0.99<br>(0.99–1.00) | 1.00<br>(1.00–1.00) | 0.99<br>(0.98–1.00) |
| Drug Use | 0.96<br>(0.95–0.98) | 0.96<br>(0.93–0.98) | 0.97<br>(0.96–0.99) | 0.97<br>(0.95–0.99) | 1.00<br>(1.00–1.00) | 0.94<br>(0.91–0.97) |
| Overall | 0.97<br>(0.95–0.97) | 0.96<br>(0.95–0.97) | 0.97<br>(0.96–0.98) | 0.97<br>(0.96–0.98) | 0.98<br>(0.95–0.99) | 0.97<br>(0.95–0.98) |

\* The overall metrics report macro-average across all SBDH categories.

## RESULTS

We included 7,225 clinical notes from 6,382 patients for method development and 971 notes from 437 patients for independent validation (Table 1). The combined patient cohort from the two MIMIC-III datasets was predominantly female (56.3%), white (72.5%), and non-Hispanic (86.1%). Similarly, the UTSW cohort was predominantly female (51.5%), white (55.1%), and non-Hispanic (79.6%). The UTSW cohort had a higher proportion of Black (33.0%) and Hispanic (14.0%) populations compared to the two MIMIC-III cohorts (Table 1). For all SBDH categories except housing, the UTSW datasets had a higher documentation rate. Additional patient characteristics are summarized in Table 1.

The distribution of human-annotated SBDH attributes across all three datasets are summarized in Figure 2 and Supplementary Figure 1. Incorporation of different SBDH terminologies in these datasets highlight the need for a generic and granular terminology to guide prompt design. In total, the original ground truths for 37 SBDH instances in the MIMIC-G and 31 instances in the MIMIC-A dataset were corrected by our team (Supplementary Table 2).

**Figure 2.**
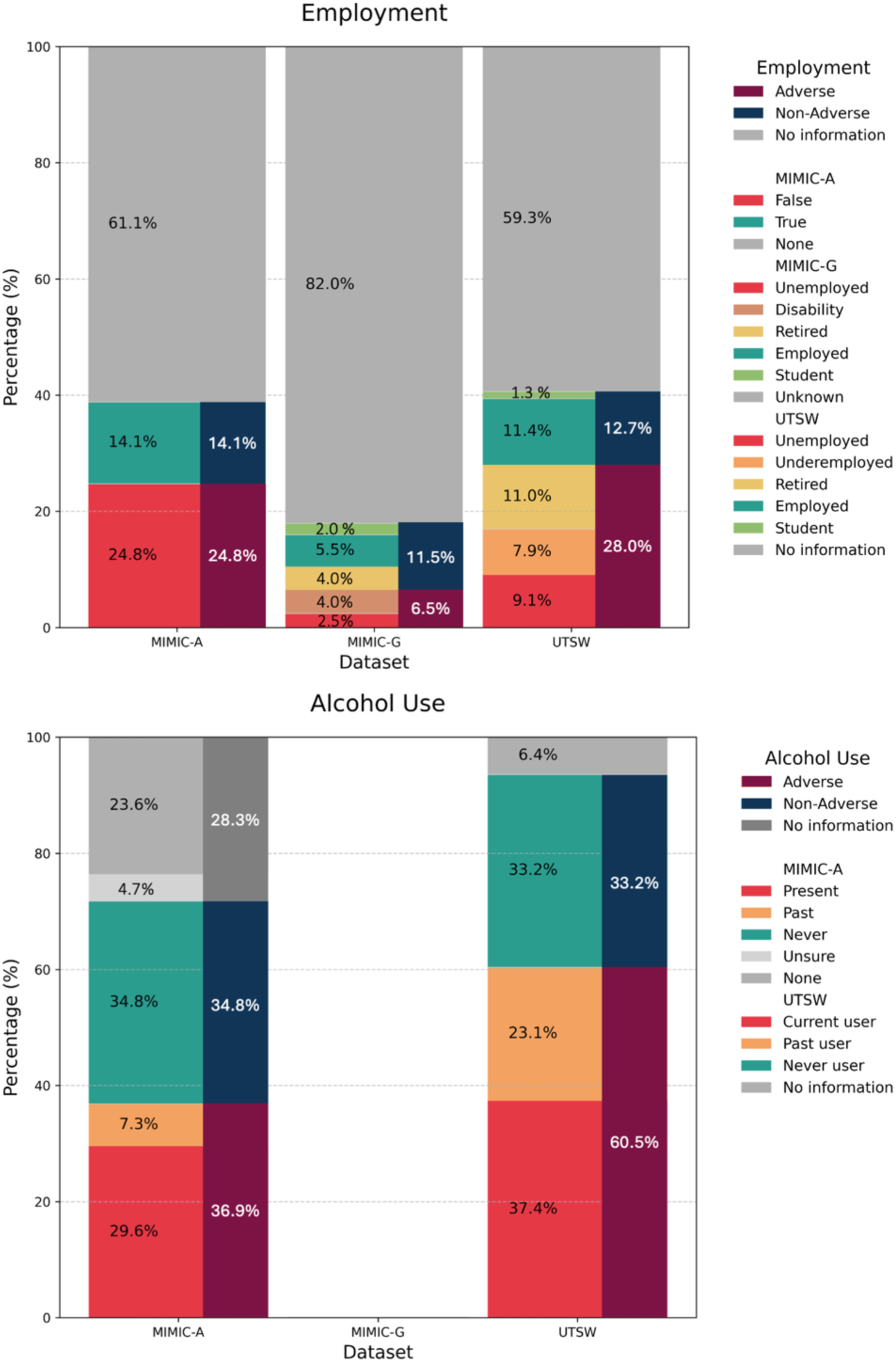
Representative SBDH attribute distribution across three datasets based on human-annotated ground truths: employment and alcohol use. In each plot, the mapping of granular attributes (left) onto the attribute classes (right)—adverse, non-adverse, or no information—is show. The original ground-truth SBDH attributes in the two MIMIC datasets differ in granularity and terminology. The terminology standard used in the MIMIC-G study assigns “retired” to the “non-adverse” class.

Through iterative prompt development using MIMIC-G and MIMIC-A, SBDH-Reader achieved high performance in extracting all SBDH categories from these two datasets (Table 3). The macro-average F1 across attributes ranged from 0.83 (MIMIC-A, employment) to 0.98 (MIMIC-G, employment). When focusing on adverse attributes, F1 ranged from 0.79 (MIMIC-A, drug use) to 1.00 (MIMIC-G, employment). For the potential clinical application of capturing adverse SBDH factors, SBDH-Reader demonstrated high sensitivity, with recall ≥ 0.87 across all categories. To extract any adverse attributes across all categories, SBDH-Reader achieved an F1 of 0.92 (MIMIC-G) and 0.89 (MIMIC-A), with recall of 0.97 (MIMIC-G) and 0.93 (MIMIC-A). Overall, lower performance was observed in MIMIC-A, with employment (macro-average F1: 0.83) and drug use (0.86) being the lowest-performing categories.

In the independent validation with the UTSW dataset, SBDH-Reader delivered an overall higher performance across all categories when compared with the development datasets (Table 3). The macro-average F1 across attributes ranged from 0.94 (employment) to 0.98 (marital relationship; tobacco use). When focusing on adverse attributes, F1 ranged from 0.96 (employment; housing) to 0.99 (tobacco use). For capturing adverse SBDH factors, SBDH-Reader demonstrated high sensitivity, with recall ≥ 0.94 across all categories, while maintaining a high precision ≥ 0.94. Performance in identifying adverse substance use (F1: 0.97–0.99) was slightly better than that extracting adverse employment, housing, and marital relationship (F1: 0.96–0.98). To extract any adverse attributes across all categories, SBDH-Reader achieved an F1 of 0.97, recall of 0.97, and precision of 0.98. Class-wise performance details are shown by confusion matrices (Figure 3). High rate of missing documentation was found for housing and employment. SBDH-Reader demonstrated consistent performance when detecting adverse and non-adverse attributes. In one particular case, SBDH-Reader classified a relatively higher proportion (25 out of 712) of missing housing documentation as non-adverse attributes. An additional test of repeatability of SBDH-Reader’s performance shows minimal variation across five independent rounds of LLM prompting (Supplementary Table 4).

**Figure 3.**
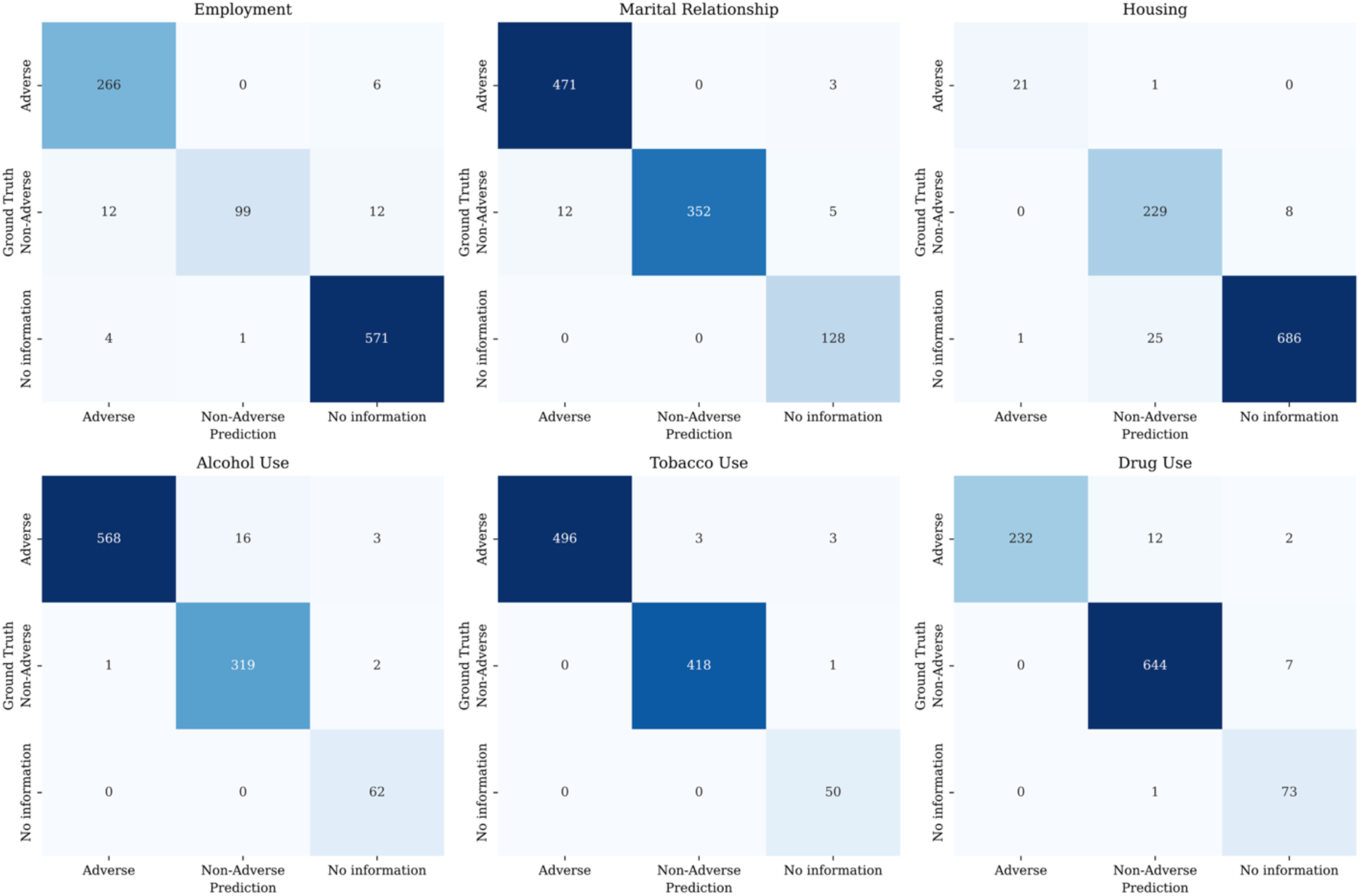
Confusion matrices for each SBDH category in UTSW validation dataset.

Through a thorough error analysis of all misclassifications in the UTSW dataset, we identified five major error types: (a) misinterpretation of the standard placeholder text, (b) misclassification despite correct evidence extraction, (c) missed supporting evidence text, (d) inference in the absence of explicit information, and (e) inference beyond the scope of the provided instructions. Detailed descriptions and potential solutions for each error type are provided in Supplementary Info 6, along with 16 examples of both correct and incorrect classifications across all SBDH categories (Supplementary Table 5). Both the quantitative (Table 3; Figure 3) and qualitative (Supplementary Info 6) evaluations highlight opportunities for further prompt engineering to improve SBDH-Reader’s performance in rare but challenging cases. The case-by-case review also uncovered instances where the original documentation was inherently ambiguous, underscoring the need to consolidate multiple note sources within a close time frame to aggregate scattered information documented by different providers, thereby enhancing completeness and clarity in real-world applications.

## DISCUSSION

The SBDH-Reader is designed to extract and classify detailed SBDH information from real-world clinical notes through direct LLM prompting. Previous NLP-based approaches for SBDH identification have been constrained by problem framing (e.g., fixed entity or relation definitions),^25,29–31^ limitations on input length due to model constraints,^25,30,31^ and the high labor costs associated with defining and annotating ground truths for model training.^24,32^ Given that SBDH documentation largely aligns with general domain knowledge and common language patterns—unlike specialized medical concepts—vertically training or fine-tuning a model for this task might not be necessary. In our study, we found that SBDH-Reader was able to accurately identify SBDH in both the development and external validation datasets, demonstrating that effective SBDH extraction can be achieved through direct LLM prompting.

One key advantage of direct LLM prompting is the ability to quickly adapt to new SBDH terminologies in a granular and flexible manner during inference. Traditionally, SBDH ontology and terminology lack widely accepted, standardized frameworks, and training models on locally defined SBDH terminologies can limit the accuracy and applicability of the results.^4,17,18^ In SBDH-Reader, the active SBDH terminology is invoked solely within the modular prompt instructions rather than embedded in the foundation model itself, allowing category- or attribute-level adjustments depending on the specific application. Additionally, SBDH-Reader generates granular SBDH attributes without preemptively reducing them into coarse, often dichotomized groups (e.g., presence vs. absence).

After SBDH-Reader extracts granular attributes, we evaluated its performance based on three post hoc aggregated classes—adverse attributes, non-adverse attributes, and no information. We included the “no information” because identifying missing SBDH documentation can serve as a care quality measure, quantifying gaps in documentation across systems, care teams, or patient populations. We reported macro-average metrics, because most SBDH categories are dominated by missing data (Table 1), leading to a highly imbalanced dataset. Reporting micro-averages in the presence of the dominant “no information” class could obscure actual performance in other classes. Additionally, we specifically reported SBDH-Reader’s performance in detecting adverse attributes due to their relevance in many clinical applications. A secondary evaluation using data subsets with all “no information” records removed for each SBDH category showed similarly high performance across all datasets (Supplementary Table 6), confirming that the metrics reported in Table 3 were not artificially inflated by the prevalence of the “non information” class. Alternatively, for specific use cases in care quality assessment—where accurately detecting missing SBDH documentation is essential—SBDH-Reader could be further refined using data with high SBDH sparsity.

Because using the MIMIC-G dataset involved aggregating the original sentence-level texts (5,328 entries)^29^ into longer, note-level texts (200 notes), we conducted a comparative analysis of running SBDH-Reader in “sentence mode” versus “note mode.” Several key differences were observed. First, 8 out of 36 employment-informing notes and 3 out of 63 marriage-informing notes contained multiple sentences with inconsistent SBDH information within the same category and note, indicating that sentence-level processing may require extra postprocessing to determine the true SBDH status for a given category. Second, the instruction section of SBDH-Reader’s prompt contains 2,412 input tokens. Processing MIMIC-G at the sentence level requires an additional 12.4 million input tokens, underscoring the cost-efficiency of operating on full notes rather than individual sentences. Third, the performance of SBDH-Reader was lower in “sentence mode” compared to “note mode” (Supplementary Table 7). This may be attributed to the high sparsity of informative content at the sentence level (missing rates: 99% for employment; 96% for marital relationship). In summary, given the ability of LLMs to handle long input texts, designing methods to process entire clinical notes offers multiple advantages over sentence-level processing.

Overall, we observed strong performance across both the development and validation datasets. Interestingly, SBDH-Reader’s performance in external validation surpassed that in the MIMIC-III development datasets. This improvement may be attributed to the more standardized SBDH terminology embedded within UTSW’s Epic system. Additionally, the supporting evidence text extracted by SBDH-Reader in the UTSW dataset was generally shorter than in the MIMIC-III datasets. This suggests that SBDH documentation in the UTSW cohort was inherently more concise, potentially due to system-wide variations.

The completeness, granularity, and accuracy of SBDH documentation in routine care are highly dependent on the healthcare system, care setting, provider type, and the time period. For example, the distribution of SBDH attributes varies substantially across the three datasets used in this study (Supplementary Figure 1). Such variations may stem from differences in patient populations between Beth Israel Deaconess Medical Center and UTSW—such as racial and ethnic backgrounds (Table 1), geographic locations, and potentially socioeconomic statuses—as well as differences in note types, associated care settings and provider types (Table 1), and data collection periods (2001–2012 vs. 2022–2023). However, for the specific goal of evaluating SBDH-Reader’s ability to extract information that is already documented in clinical notes, these variations are unlikely to negatively affect performance. On the contrary, utilization of three diverse datasets highlights SBDH-Reader’s generalizability across different patient populations, care settings, and time periods. Future studies may benefit from extending the evaluation of SBDH-Reader to other disease-specific cohorts, healthcare systems, and documentation patterns.

### Limitations and Future Directions

In this initial version of SBDH-Reader, we focused on six SBDH categories that represent a relatively broad spectrum of SBDH coverage, selected primarily based on the availability of high-quality annotated data from MIMIC and UTSW. The modular design of SBDH-Reader’s prompt structure enables future expansion onto additional SBDH categories. For example, additional socioeconomic characteristics beyond employment status and more nuanced aspects of social or community support beyond marital relationships may be of interest, although such information may be inconsistently or infrequently documented in routine clinical care.

Our results demonstrated minimal variation when re-prompting GPT-4o (Supplementary Table 4), indicating its stability. Future evaluations can be conducted as newer versions of GPTs or other advanced LLMs become available in regulatory-compliant environments. Since SBDH-Reader does not rely on model fine-tuning, validating it for new use cases or with different LLMs remains straightforward and efficient. In addition, SBDH-Reader is API based, enabling easier integration into existing care systems and workflows compared with hosting an LLM locally.

The current version of SBDH-Reader can process long sections of clinical text, up to the prompt length limit of the underlying LLM. This provides a significant advantage over earlier methods that focused on sentence- or paragraph-level texts.^25,30,31^ For example, GPT-4o can handle up to 128k tokens, or approximately 170,000 English words, which is sufficient for typical clinical notes. However, future iterations could extend the scope by incorporating longer sequences of notes for more comprehensive and longitudinal SBDH information collection. For example, in this study, while we included multiple notes for some patients (Table 1), we focused on evaluating the SBDH-Reader’s performance in each individual note without considering potential changes in patient’s status across different time points of note-taking. The current prompt specifically instructs the LLM to extract the patient’s current SBDH status at the time of the note (Supplementary Info 3). One potential limitation of this design is that it may overlook historical SBDH information documented within the same note, if present. This limitation may be mitigated in the future versions of SBDH-Reader by incorporating the aforementioned longitudinal SBDH extraction capability.

SBDH reported by individual providers may include incomplete or inaccurate information, as well as implicit bias. In addition, given the substantial amount of missing SBDH documentation in real-world clinical notes, investigating how to properly interpret and handle the implications of such missingness is important. For SBDH-Reader to achieve real-world impact, future studies should explore how to effectively integrate and consolidate available notes, structured EHR data, and area-level SBDH measures into comprehensive patient-level SBDH profiles. During large-scale deployment, the overall cost burden may become a concern depending on data volume and system constraints. Further investigations into which note types and authors yield more accurate and abundant SBDH documentation could help optimize processing priorities and reduce costs. Since most clinical notes only dedicate a few sections to SBDH information, a method for effectively trimming full-length notes to focus on these sections could improve efficiency.

## CONCLUSION

We developed SBDH-Reader, an LLM-powered method for extracting note-level, categorized, and granular SBDH information from real-world clinical notes. Through effective prompt engineering, the SBDH-Reader demonstrated high performance in extracting and classifying SBDH data across six key categories. While additional validation across diverse health systems, care settings, and patient populations is needed, the SBDH-Reader shows potential as an effective tool for collecting real-world SBDH data to support clinical research and patient care.

## CONTRIBUTORSHIP

Zifan Gu (Conceptualization, Data curation, Formal analysis, Investigation, Methodology, Software, Validation, Visualization, Writing - original draft, Writing - review & editing); Lesi He (Data curation, Formal analysis, Investigation, Methodology, Software, Validation, Visualization, Writing - original draft, Writing - review & editing); Awais Naeem (Data curation, Formal analysis, Investigation, Methodology, Software, Validation, Visualization); Pui Man Chan (Data curation, Formal analysis, Writing - review & editing); Asim Mohamed (Data curation); Hafsa Khalil (Data curation); Yujia Guo (Data curation); Jingwei Huang (Methodology); Ismael Villanueva-Miranda (Methodology); Ying Ding (Resources, Supervision); Wenqi Shi (Methodology, Writing - review & editing); Matthew E. Dupre (Conceptualization, Investigation, Methodology, Writing - review & editing); Guanghua Xiao (Funding acquisition, Investigation, Methodology, Resources, Supervision, Writing - review & editing); Eric D. Peterson (Conceptualization, Investigation, Methodology, Resources, Supervision, Writing - review & editing); Yang Xie (Conceptualization, Funding acquisition, Investigation, Methodology, Resources, Supervision, Writing - review & editing); Ann Marie Navar (Conceptualization, Investigation, Methodology, Resources, Supervision, Writing - original draft, Writing - review & editing); Donghan M. Yang (Conceptualization, Data curation, Formal analysis, Funding acquisition, Investigation, Methodology, Resources, Software, Supervision, Validation, Visualization, Writing - original draft, Writing - review & editing)

## DATA AVAILABILITY

The MIMIC-III data are available to credentialed users at https://physionet.org. The corrected ground truths generated from this study for the two published MIMIC-III datasets will be made available on PhysioNet. The UTSW dataset is not publicly available due to data and privacy protection policies at UTSW.

## Supplementary Info 1. Note selection process for the UTSW validation dataset

To select notes with more abundant SBDH documentation for evaluating SBDH-Reader’s performance, we conducted a keyword search within the “Social History” section of all notes available from 2022 to 2023 for the heart failure cohort at UTSW. The keywords used in this process included the most frequently seen terms describing the six SBDH categories, based on our manual review. For each category, we randomly selected notes containing the category-specific keywords to ensure that at least 30 notes were included for each attribute of that category. This process was iterated for all categories in the following order: housing, marital relationship, employment, drug use, alcohol use, and tobacco use. The entire process resulted in a dataset containing 971 notes for 437 patients, with a lower missing rate compared to the other two MIMIC-III datasets (Table 1).

## Supplementary Info 2. SBDH annotation guidelines

### Terminology

- “Category”: A specific SBDH domain or factor. The current guidelines cover six SBDH Categories: employment, housing, marital relationship, alcohol use, tobacco use, and drug use. A given input text may contain descriptions of multiple Categories and may also include multiple descriptions within a single Category.
- “Attribute”: A pre-defined value that characterizes a given SBDH category. For example, for the Category of “Marital Relationship”, permissible Attributes include married, partnered, widowed, divorced, other singleness, and other (see detailed definitions in Table 2). The permissible Attributes are mutually exclusive. Each SBDH Category may be assigned only one Attribute at a given time.
- “Instance”: Any single piece of evidence used to assign an Attribute to a given SBDH Category is considered one Instance. A single input text may contain multiple Instances related to the same SBDH Category, and these Instances may support different Attributes. The annotation task involves: (a) identifying and recording all Instances that describe the Category, (b) consolidating the information from all Instances to determine one Attribute that best represents the <u>current status of</u> <u>the Category</u> at the time the note was written, and (c) assigning that final, selected Attribute to the Category.

### Resources

- Category and Attribute definition: Table 2
- Example annotation:

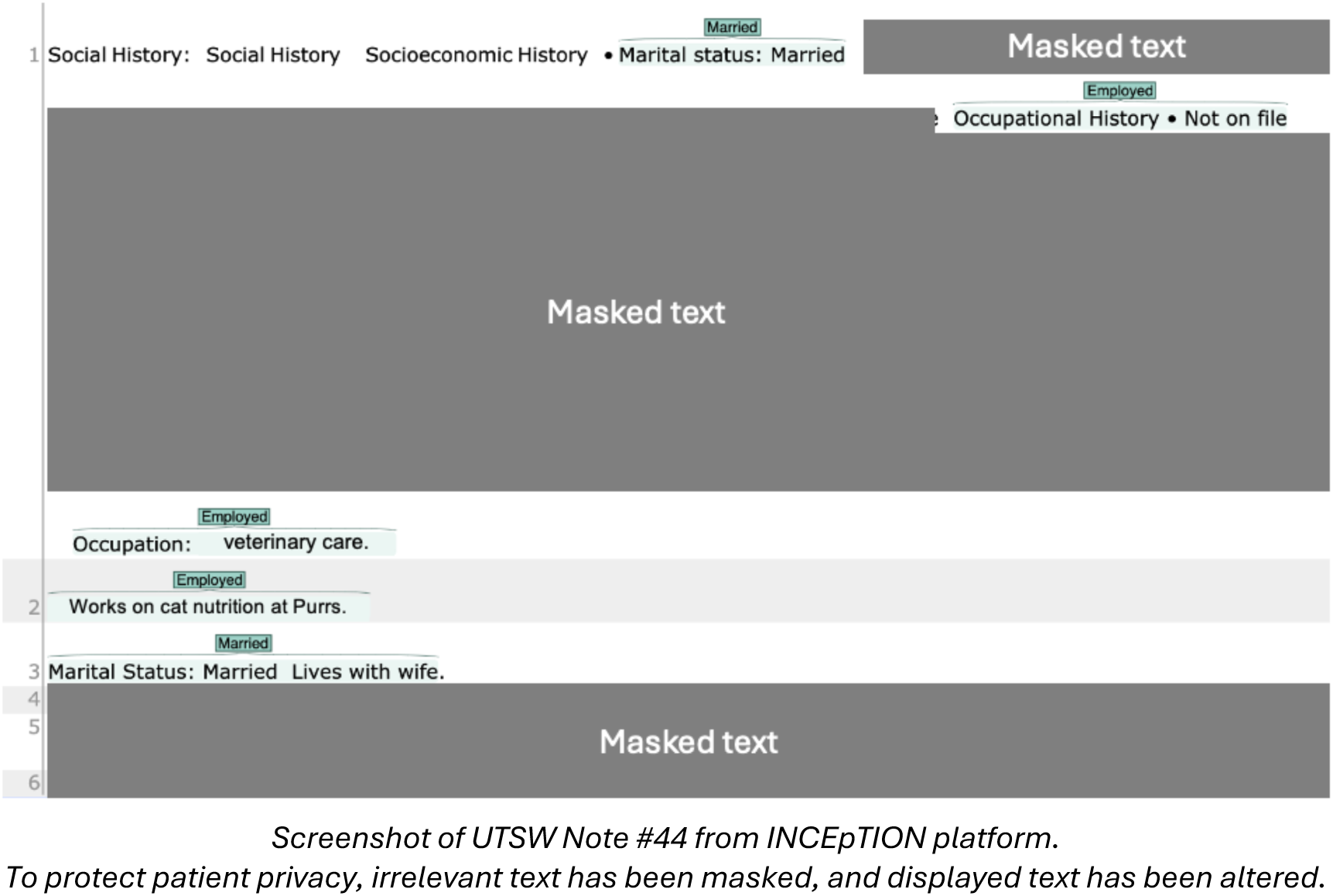
Screenshot of UTSW Note #44 from INCEpTION platform. To protect patient privacy, irrelevant text has been masked, and displayed text has been altered.

### Guidelines

- If no Instance can be found for a Category in a note, please do not label anything for this Category and proceed to the next Category or note.
- Only label “No information” for a Category if the note states “Not on file”, “No information” etc. for that Category (Example: Marital Status: Not on file)
- When multiple Instances are present in a note, highlight each of them, and assign them the same Attribute after summarizing the info from all these Instances. If conflicts occur between Instances, try to resolve and assign the same (final) Attribute.

In the example above (Note #44):

- There are 2 Instances for Marital Relationship, please label both instances.
- There are 3 Instances for Employment with conflicts between them. Please try to decide on the final Attribute based on the whole note and label all Instances with the final Attribute.

**Supplementary Table 1.** Inter-annotator agreement for the first 100 notes in UTSW dataset.

| Category | Cohen's Kappa | Percent Agreement |
| --- | --- | --- |
| Employment | 0.97 | 98% |
| Housing | 0.85 | 98% |
| Marital Relationship | 0.97 | 98% |
| Alcohol Use | 0.97 | 98% |
| Tobacco Use | 0.98 | 99% |
| Drug Use | 0.98 | 99% |

## Supplementary Info 3. Final prompt and example output

Prompt structure is outlined (Figure 1). For privacy protection, the example outputs for various SBDH categories were extracted from different patients.

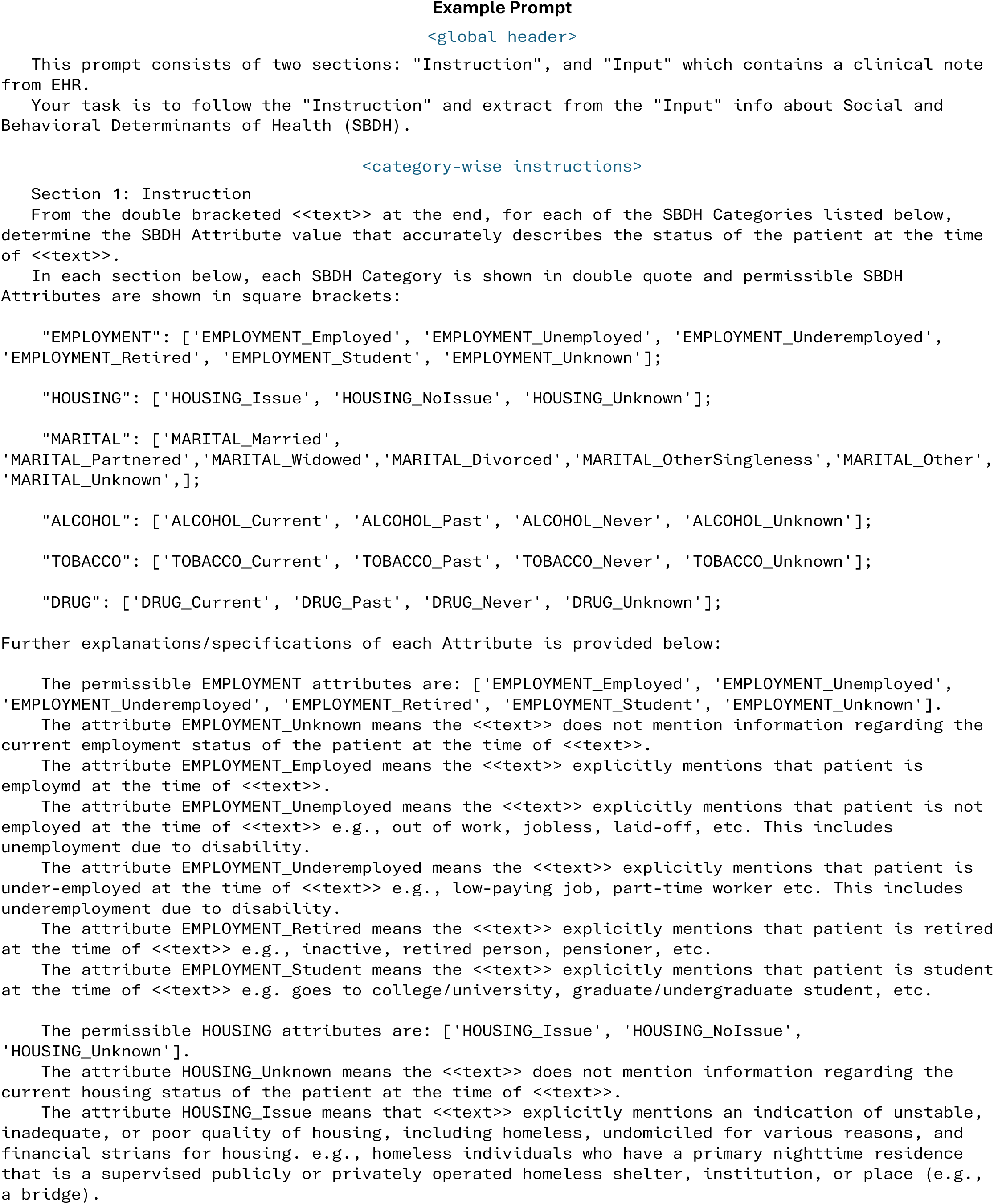
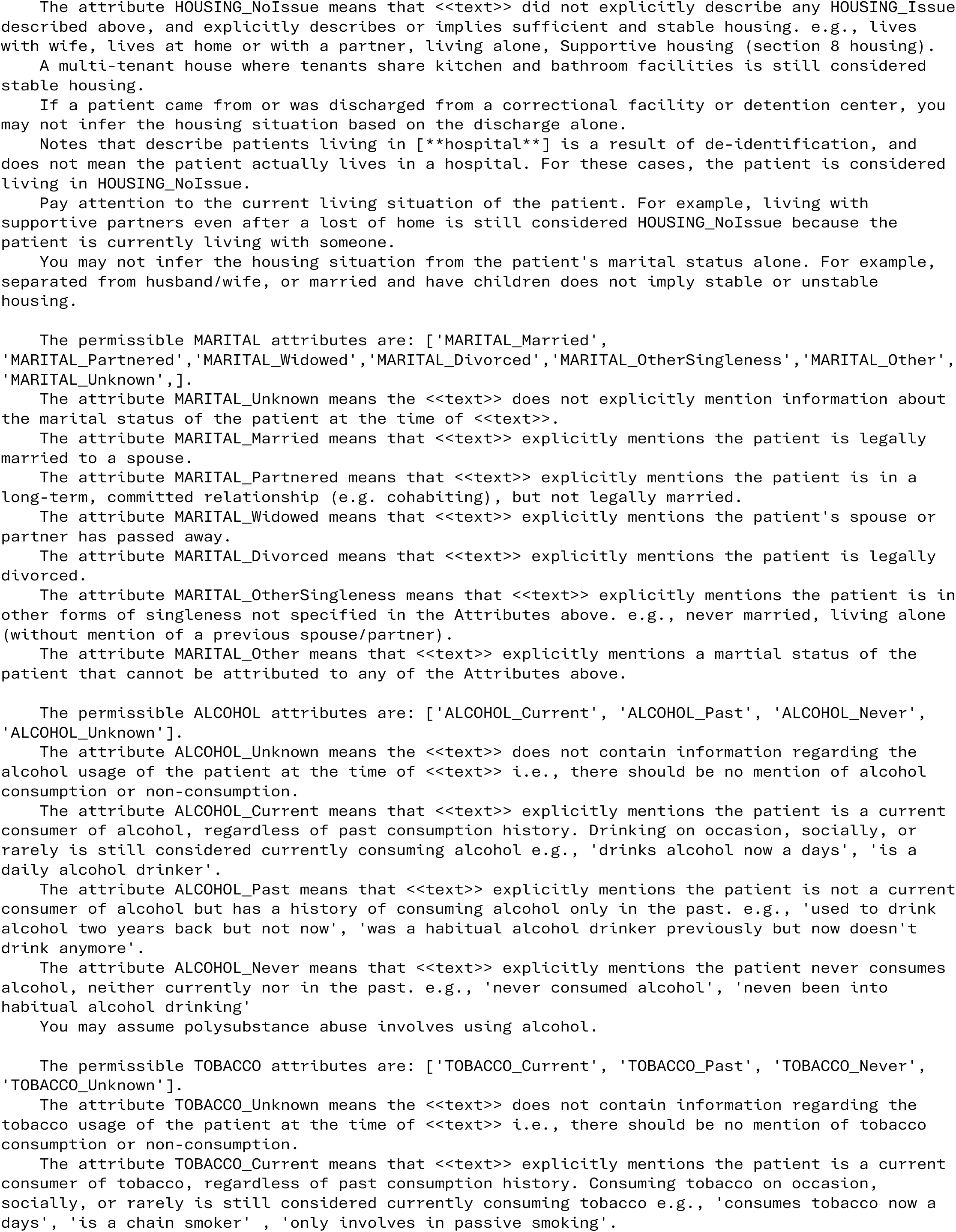
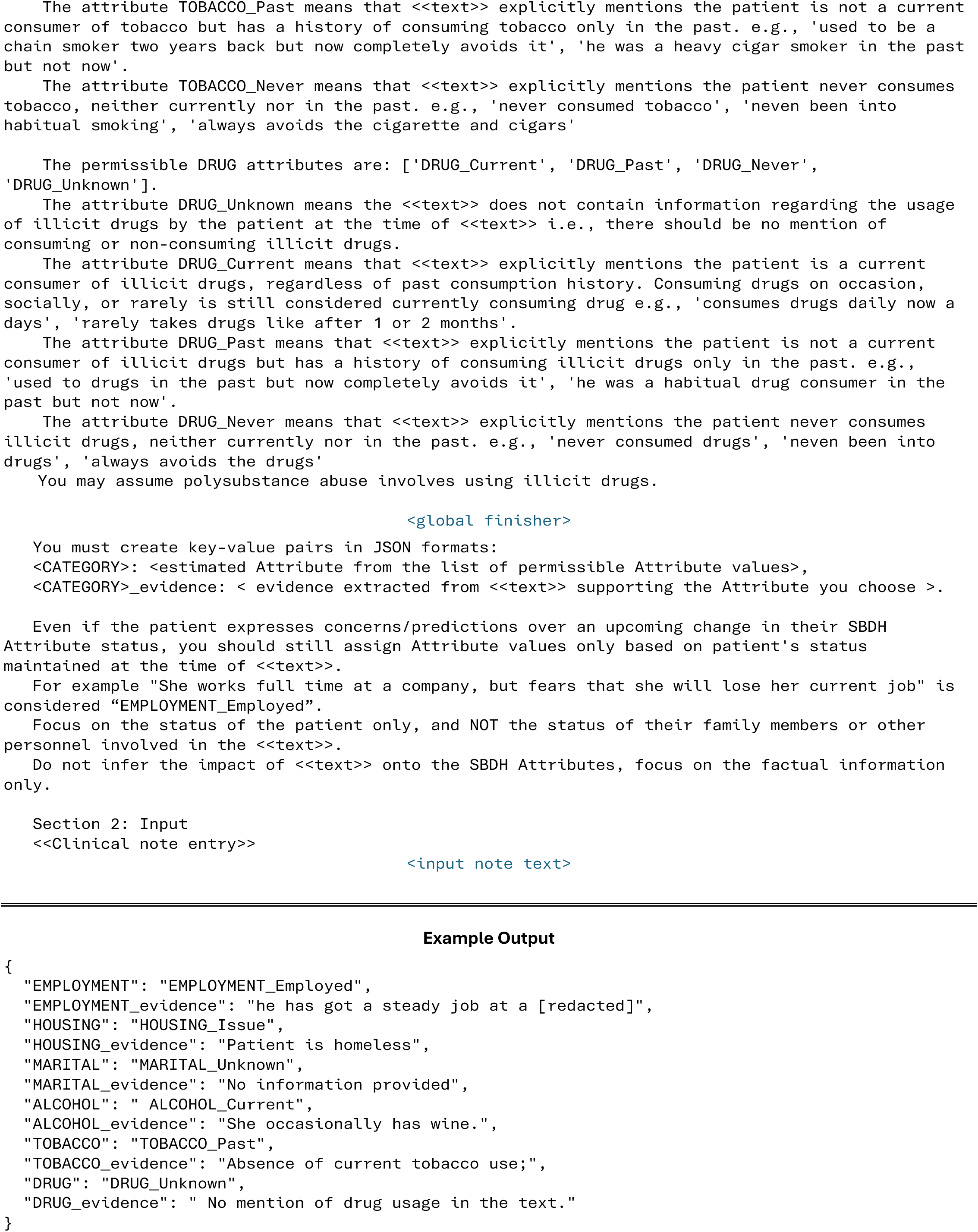

## Supplementary Info 4. Milestone prompt versions throughout the iterative prompt refinement process

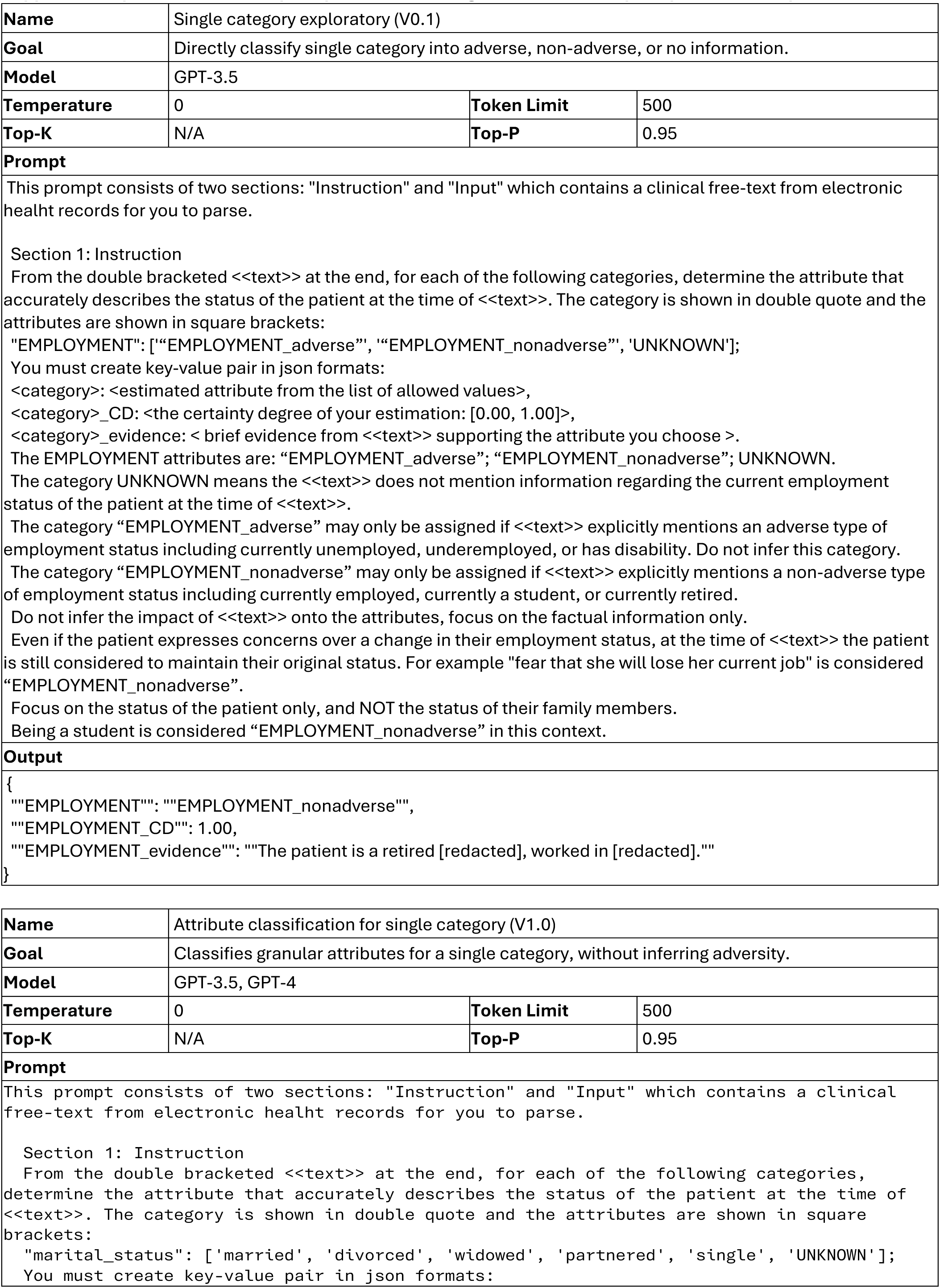
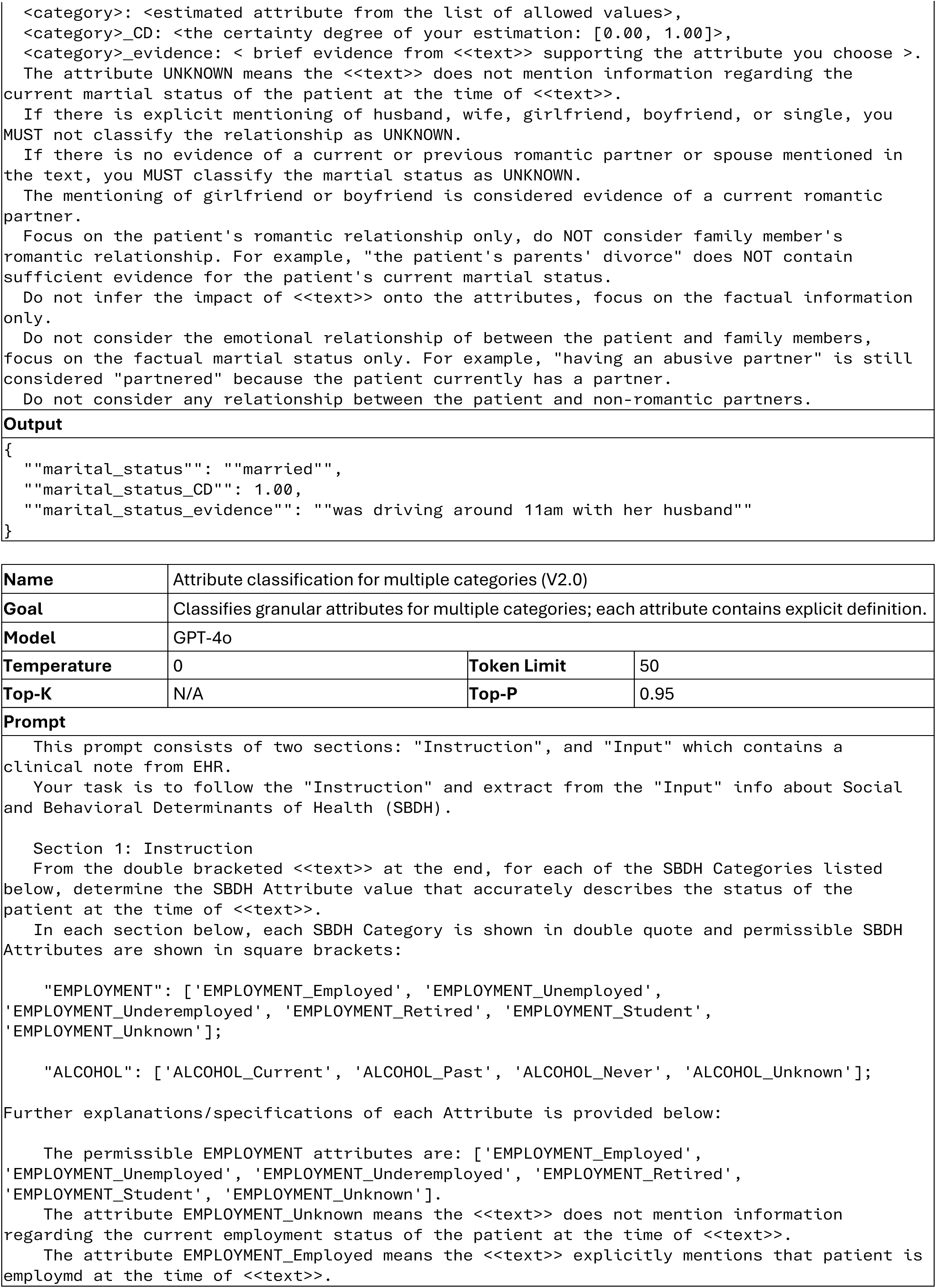
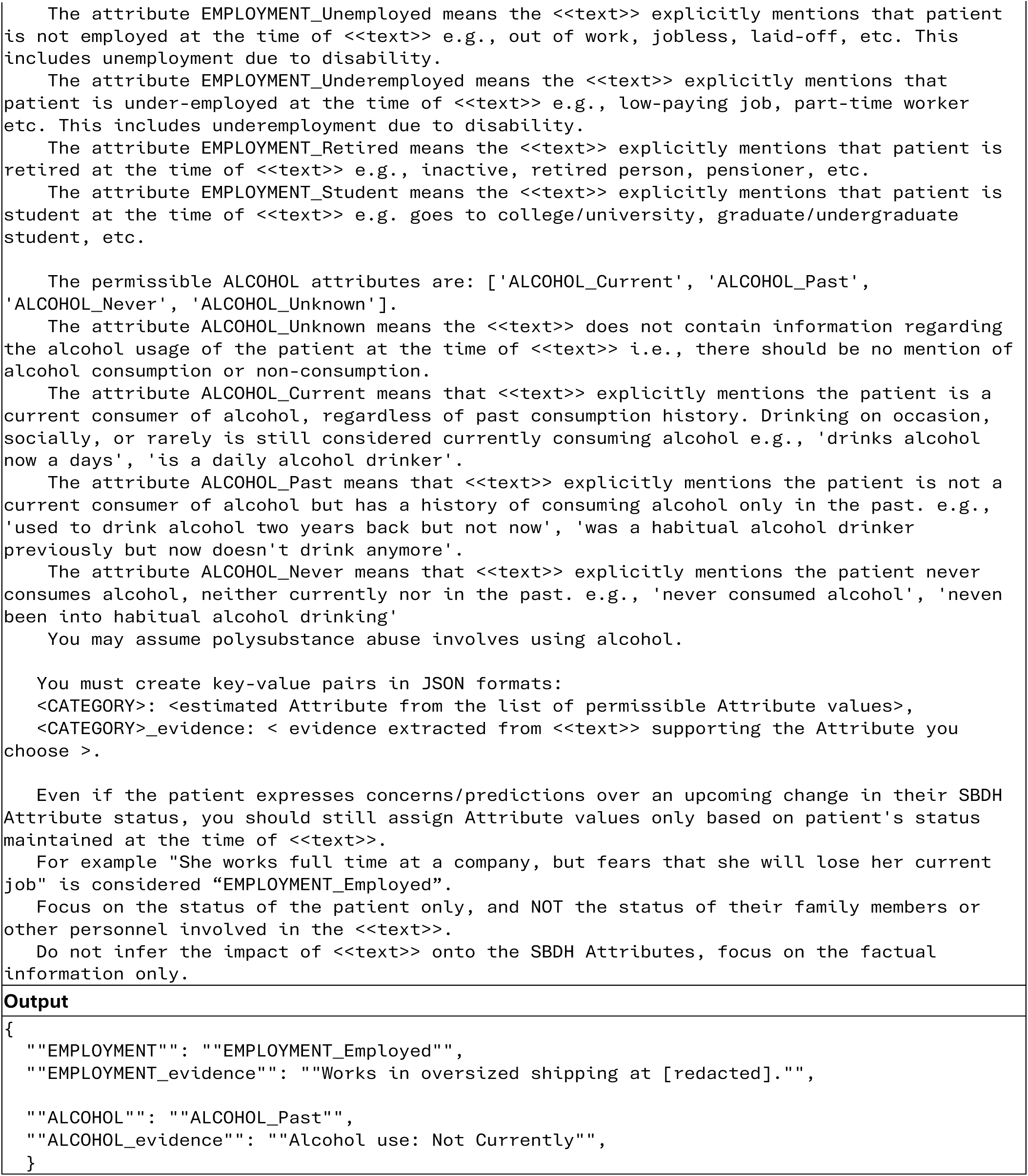

## Supplementary Info 5. Iterative process of prompt refinement accompanied by ground truth review and correction in the MIMIC-G and MIMIC-A datasets

To develop and refine the LLM prompt using MIMIC-G and MIMIC-A data, we iteratively added and modified specifications for each SBDH category and attribute present in these two development datasets. For each low-performing attribute, we compared the original notes with the supporting evidence text extracted by the LLM. If misclassifications made by the LLM were found to be repeatable across multiple notes—indicating systematic rather than isolated errors—we incorporated new specifications into the prompt to prevent such errors. For example, some notes describe patients as “living in [**hospital**],” a placeholder resulting from MIMIC-III’s de-identification protocol. The LLM mistakenly interpreted this as unstable housing (i.e., that the patient was literally living in a hospital). We resolved this issue by adding a prompt instruction that such syntax patterns are artifacts of de-identification.

We began the prompt refinement process with MIMIC-G, as it contains ground truths with more granular attributes. The iterative refinement continued until minimal improvement was observed from one iteration to the next—typically when remaining errors were case-specific and not generalizable. The entire MIMIC-G dataset was involved. We then proceeded to MIMIC-A, which has less granular ground truths (e.g., only three employment attributes compared to six in MIMIC-G; see Supplementary Figure 1). Due to its large size, we first focused on a randomly selected 20% subset of MIMIC-A, and after the general performance was acceptable, we proceeded to the entire dataset to review the remaining errors. We followed the same procedure of analyzing underperforming attributes, updating prompt specifications, and concluding the process upon observing diminishing returns.

During this manual error analysis, we identified several cases where the LLM made correct classifications with valid supporting evidence, but the original ground truth labels from MIMIC-G or MIMIC-A were incorrect (see examples in Supplementary Table 3). In such cases, after team review and discussion (ZG, LH, DMY), we documented the note ID, the original ground truth, and the proposed revised ground truth in a tracking table. This process resulted in three rounds of ground truth revision for MIMIC-A and two rounds for MIMIC-G. In total, 37 instances in MIMIC-G and 31 instances in MIMIC-A were corrected (Supplementary Table 2). We are currently in the process of releasing these corrected ground truths via PhysioNet, in accordance with MIMIC-III’s policies and standards.

**Supplementary Table 2.**
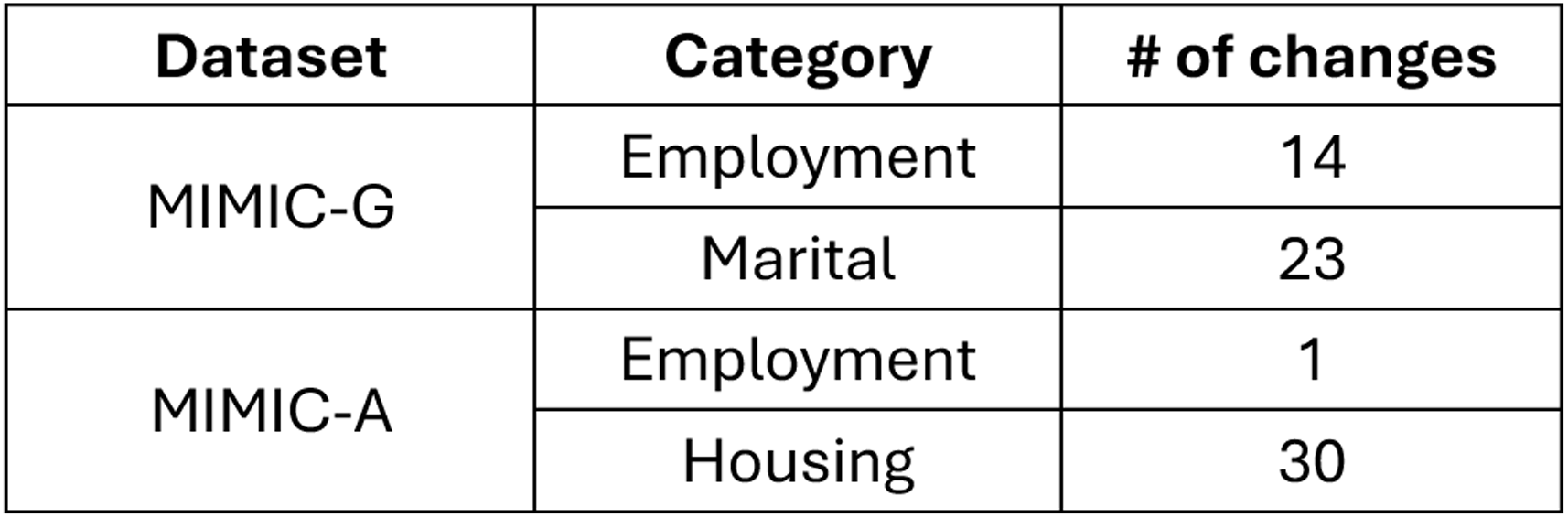
Summary of ground truth corrections in the MIMIC-G and MIMIC-A datasets.

**Supplementary Table 3.**
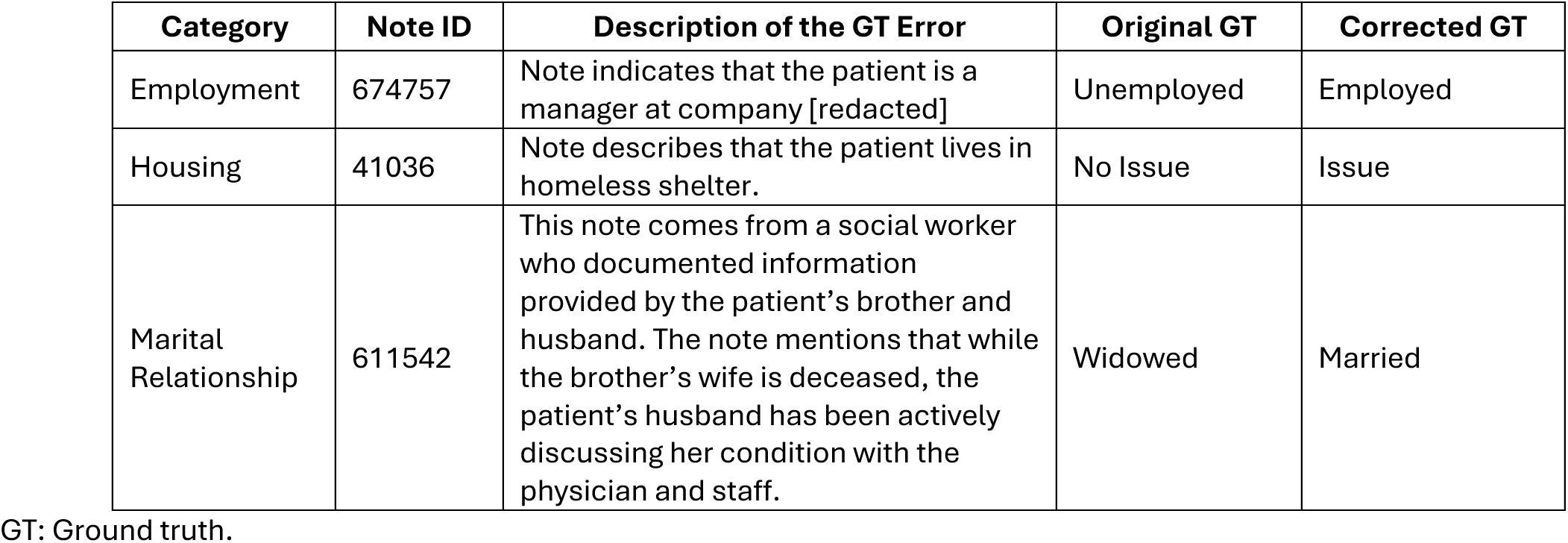
Examples of corrected ground truths in the MIMIC-G and MIMIC-A datasets.

| Category | Note ID | Description of the GT Error | Original GT | Corrected GT |
| --- | --- | --- | --- | --- |
| Employment | 674757 | Note indicates that the patient is a manager at company [redacted] | Unemployed | Employed |
| Housing | 41036 | Note describes that the patient lives in homeless shelter. | No Issue | Issue |
| Marital Relationship | 611542 | This note comes from a social worker who documented information provided by the patient’s brother and husband. The note mentions that while the brother’s wife is deceased, the patient’s husband has been actively discussing her condition with the physician and staff. | Widowed | Married |

GT: Ground truth.

**Supplementary Figure 1.**
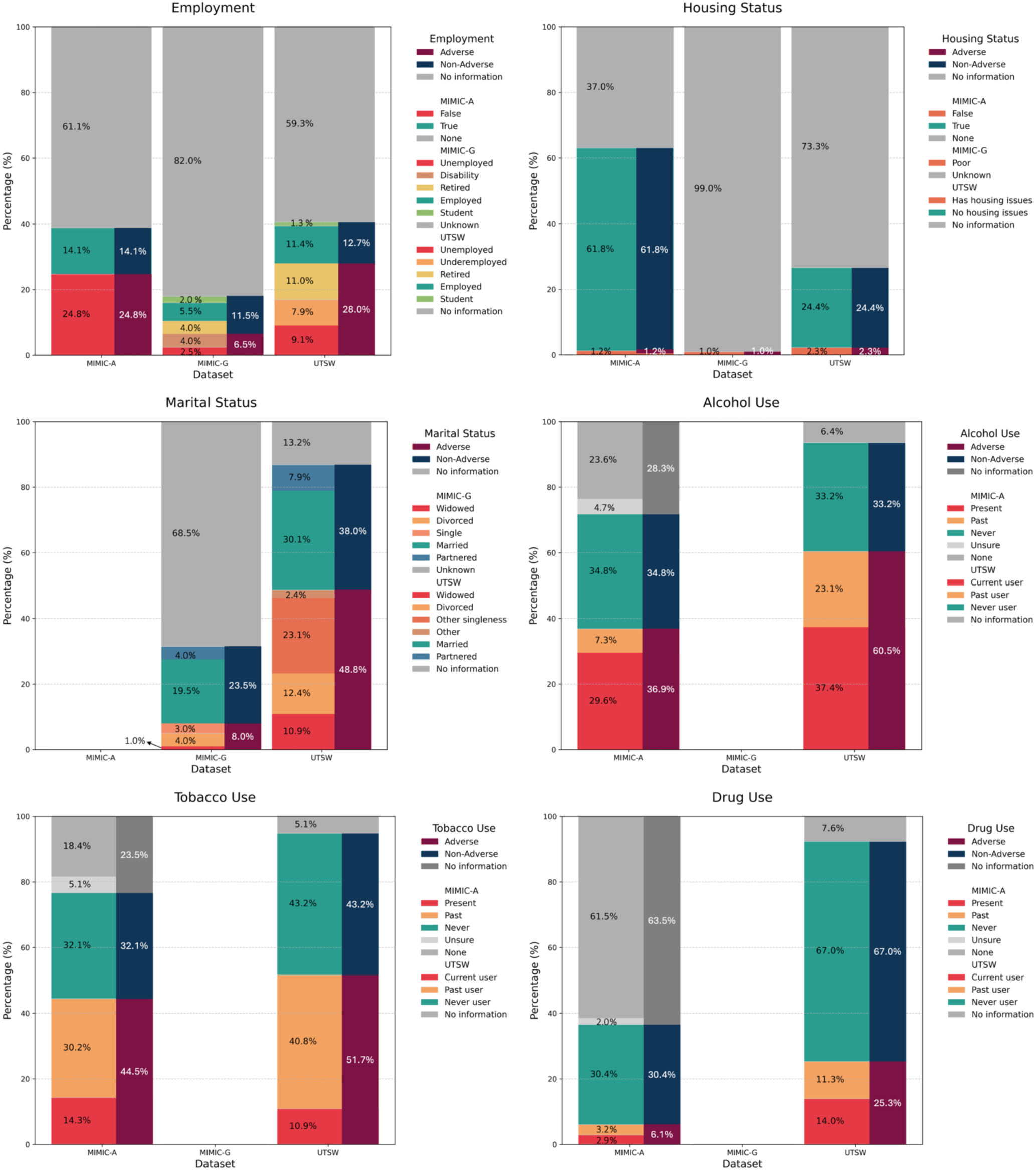
Summary of SBDH attribute distribution across three datasets based on human-annotated ground truths. In each plot, the mapping of granular attributes (left) onto the attribute classes (right)—adverse, non-adverse, or no information—is show. The original ground-truth SBDH attributes in the two MIMIC datasets differ in granularity and terminology. The terminology standard used in the MIMIC-G study assigns “retired” to the “non-adverse” class.

**Supplementary Table 4.** SBDH-Reader’s performance variation across five independent rounds of LLM prompting. The median and standard deviation are estimated from five independent promptings using GPT-4o.

|  | Macro average |  |  | Adverse attributes only |  |  |
| --- | --- | --- | --- | --- | --- | --- |
| Category | F1 | Precision | Recall | F1 | Precision | Recall |
| MIMIC-G |  |  |  |  |  |  |
| Employment | 0.98 ± 0.00 | 0.96 ± 0.00 | 0.99 ± 0.00 | 1.00 ± 0.00 | 1.00 ± 0.00 | 1.00 ± 0.00 |
| Marital Relationship | 0.90 ± 0.01 | 0.89 ± 0.00 | 0.91 ± 0.02 | 0.83 ± 0.01 | 0.75 ± 0.00 | 0.94 ± 0.03 |
| MIMIC-A |  |  |  |  |  |  |
| Employment | 0.83 ± 0.00 | 0.81 ± 0.01 | 0.86 ± 0.00 | 0.84 ± 0.00 | 0.80 ± 0.00 | 0.87 ± 0.00 |
| Housing | 0.90 ± 0.00 | 0.89 ± 0.01 | 0.91 ± 0.00 | 0.85 ± 0.01 | 0.79 ± 0.02 | 0.92 ± 0.00 |
| Alcohol use | 0.91 ± 0.00 | 0.93 ± 0.00 | 0.90 ± 0.00 | 0.93 ± 0.00 | 0.89 ± 0.00 | 0.98 ± 0.00 |
| Tobacco use | 0.94 ± 0.00 | 0.95 ± 0.01 | 0.93 ± 0.00 | 0.97 ± 0.00 | 0.95 ± 0.00 | 0.99 ± 0.00 |
| Drug use | 0.88 ± 0.00 | 0.86 ± 0.00 | 0.91 ± 0.00 | 0.82 ± 0.01 | 0.77 ± 0.00 | 0.87 ± 0.00 |
| UTSW |  |  |  |  |  |  |
| Employment | 0.94 ± 0.00 | 0.97 ± 0.00 | 0.96 ± 0.00 | 0.95 ± 0.01 | 0.95 ± 0.02 | 0.95 ± 0.00 |
| Housing | 0.96 ± 0.01 | 0.95 ± 0.00 | 0.92 ± 0.00 | 0.96 ± 0.00 | 0.94 ± 0.00 | 0.98 ± 0.00 |
| Marital Relationship | 0.98 ± 0.00 | 0.97 ± 0.00 | 0.96 ± 0.00 | 0.95 ± 0.01 | 0.95 ± 0.02 | 0.95 ± 0.00 |
| Alcohol use | 0.97 ± 0.00 | 0.96 ± 0.00 | 0.99 ± 0.0 | 0.98 ± 0.00 | 1.00 ± 0.00 | 0.97 ± 0.00 |
| Tobacco use | 0.98 ± 0.00 | 0.97 ± 0.00 | 1.00 ± 0.00 | 0.99 ± 0.00 | 1.00 ± 0.00 | 0.99 ± 0.00 |
| Drug use | 0.96 ± 0.00 | 0.96 ± 0.00 | 0.97 ± 0.00 | 0.97 ± 0.01 | 1.00 ± 0.00 | 0.94 ± 0.01 |

## Supplementary Info 6. Error analysis

The error analysis conducted in this study is based on evaluating the outputs generated by the final version of SBDH-Reader— developed through iterative prompt engineering using the two MIMIC-III datasets—against human-annotated ground truths from the independent UTSW testing dataset.

The quantitative error analysis is presented in Table 3 and Figure 3, and is discussed in detail in the Results and Discussion sections. In this Supplementary Information section, we provide a qualitative error analysis by characterizing key error patterns through a case-by-case review of all misclassifications made by SBDH-Reader on the UTSW dataset. Representative examples of both correct and incorrect classifications for each SBDH category are included in Supplementary Table 5, indexed by Case ID.

### Error Type A. Over-attention to the standard “Not On File” placeholder text and neglect of other relevant information

#### Example(s)

Cases 3, 8

#### Description

In the UTSW Epic system, the auto-populated placeholder text “Not On File” frequently appears in each SBDH section by default if the provider does not manually overwrite it. Depending on individual documentation habits, providers may leave this standard text in place even after adding relevant SBDH information in subsequent narrative sections. When both “Not On File” and additional SBDH details co-exist, SBDH-Reader often prioritizes the placeholder and incorrectly classifies the note as “unknown” (no information). For example, a patient’s occupation was recorded as “Not On File,” yet another section discussing stressors included narrative text citing concerns related to “work, family, and church” (Case 3). Similarly, a patient’s marital status was marked as “Not On File,” while narrative notes indicated the patient “lives with wife” (Case 8).

#### Potential Solution

Introduce a healthcare system-specific header in the prompt to instruct the LLM to consider relevant narrative descriptions beyond standard placeholder text such as “Not On File.” This customization may need to reflect the EHR system’s auto-population behavior.

### Error Type B. Misclassification despite correct evidence extraction

#### Example(s)

Cases 2, 11, 12, 16

#### Description

SBDH-Reader accurately extracted the supporting evidence text but still assigned an incorrect attribute. For example, when “Occupation” was documented but “Employer” was listed as “NONE,” followed by the actual company name mentioned in the “Comment” field, the model incorrectly inferred unemployment instead of recognizing the missing “Employer” entry as a non-informative absence (Case 2). Similarly, for alcohol use, SBDH-Reader correctly identified text indicating that the patient denies any alcohol consumption, yet it misclassified the case as “Past User” (Case 11).

#### Potential Solution

Enhance the prompt with more precise specifications to consolidate multiple instances of supporting evidence. Incorporate few-shot examples illustrating subtle language cues associated with each classification to improve LLM interpretation and reduce this error pattern.

### Error Type C. Missed supporting evidence text

#### Example(s)

Case 9, 14

#### Description

SBDH-Reader failed to extract the relevant supporting evidence text that is present in the note. This results in either (a) no evidence being extracted at all, or (b) irrelevant text being selected as evidence. As a result, the assigned attribute was wrong. For example, one note explicitly described the patient as a former tobacco user, with detailed descriptions of packs/day and pack year, yet the LLM incorrectly reported that there was no information regarding the patient’s tobacco use status (Case 14). In another example, documentation of the patient’s marital status and spouse’s name was omitted by the LLM (Case 9).

#### Potential Solution

Strengthen the prompt with additional guidance emphasizing complete context scanning. Incorporating more varied few-shot examples, especially those with subtle or embedded SBDH clues.

### Error Type D. Inference in the absence of explicit information

#### Example(s)

Case 6

#### Description

SBDH-Reader failed to distinguish between informative and non-informative absences. For instance, while the notes contained no mention of a housing situation, the LLM still assigns a label of Housing: No Issue, rather than acknowledging the lack of information (Case 6).

#### Potential Solution

Add a global instruction to distinguish lack of information and non-adverse status.

### Error Type E. Inferring beyond the scope of the provided instructions

#### Example(s)

Case 5

#### Description

SBDH-Reader made inferences that extended beyond the scope of prompt instructions. While reasonable inference is sometimes necessary, the LLM drew conclusions that were not directly supported by the text. For example, the patient reported financial strain as difficulty paying living expenses, and the LLM inferred housing issues (Case 5). While housing issues may arise in the presence of significant financial strain, the LLM should not infer such issues unless they are directly described in relation to housing.

#### Potential Solution

Add a global instruction to limit inference to information directly related to the specific SBDH category, avoiding assumptions based on indirect or unrelated descriptions.

### Distribution across major error types

We observed that the distribution of error types varied across SBDH categories. A summary is provided in the following table.

| Category | Total | Type A | Type B | Type C | Type D | Type E | Other |
| --- | --- | --- | --- | --- | --- | --- | --- |
| Employment | 35 | 2 | 9 | 5 | 10 | 4 | 5 |
| Housing | 35 | 4 |  | 2 | 3 | 17 | 9 |
| Marital Relationship | 20 | 18 |  | 2 |  |  | 0 |
| Alcohol Use | 22 | 8 | 2 | 5 |  |  | 7 |
| Tobacco Use | 7 | 2 | 1 | 4 |  |  | 0 |
| Drug Use | 22 | 6 | 8 | 7 |  |  | 1 |

### Other Cases

Besides the five common error types listed above, we also identified several rare but important insights from the error analysis. We found cases involving inherent ambiguity, where even human annotators may reasonably differ in interpretation. For example, one note lists the occupation as “disability,” while a free-text comment states “electrical engineer.” It is unclear—and equally plausible—that the patient was formerly an electrical engineer and is now disabled, or that they are currently employed as an electrical engineer with appropriate accommodations. In another case, alcohol use is labeled as “No,” with a comment stating “none in the last month.” This could reflect either: (1) the patient does not drink at all and is a never user, or (2) the patient does drink alcohol but has not consumed any in the past month, indicating a past user. Such examples highlight the challenges of inferring intent and context, even when explicit text is available. Such scenarios highlight not merely the limitations of either LLMs or human annotators, but rather the limitations in the quality of SBDH documentation during routine clinical care. Future work may benefit from consolidating multiple note sources within a close time frame to aggregate scattered information documented by various providers, thereby achieving greater completeness and clarity.

The housing category presented unique challenges compared to the other SBDH categories. Notably, it was the only category in which we observed a hallucination. SBDH-Reader supported a “Housing: No Issue” classification with the statement “Patient was diuresed and discharged home,” despite no mention in the text of discharge disposition. We also identified a case of error propagation in the housing category—again, the only such instance across all SBDH categories. In this case, the SBDH-Reader incorrectly extracted the phrase “accompanied by his wife” and inferred stable housing, even though the note explicitly documented housing instability, including the patient’s report of being unable to pay for housing in the past year. It is unclear why these rare errors were concentrated in the housing domain. Future work should therefore target prompting strategies to mitigate these infrequent yet impactful errors.

**Supplementary Table 5.**
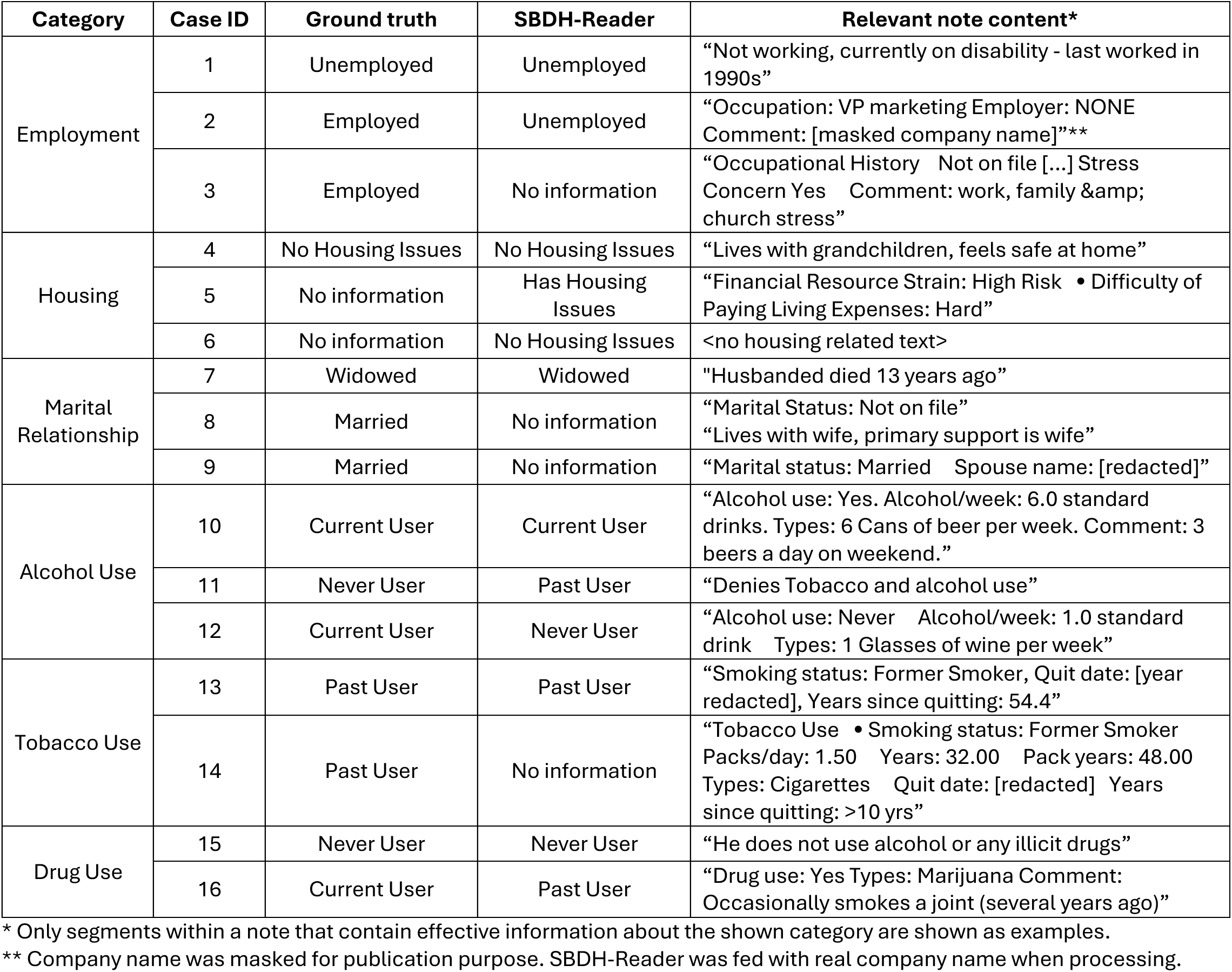
Examples of SBDH-Reader outputs and error analysis in the UTSW dataset.

| Category | Case ID | Ground truth | SBDH-Reader | Relevant note content* |
| --- | --- | --- | --- | --- |
| Employment | 1 | Unemployed | Unemployed | "Not working, currently on disability - last worked in 1990s" |
|  | 2 | Employed | Unemployed | "Occupation: VP marketing Employer: NONE Comment: [masked company name]"** |
|  | 3 | Employed | No information | "Occupational History Not on file [...] Stress Concern Yes Comment: work, family & church stress" |
| Housing | 4 | No Housing Issues | No Housing Issues | "Lives with grandchildren, feels safe at home" |
|  | 5 | No information | Has Housing Issues | "Financial Resource Strain: High Risk • Difficulty of Paying Living Expenses: Hard" |
|  | 6 | No information | No Housing Issues | <no housing related text> |
| Marital Relationship | 7 | Widowed | Widowed | "Husband died 13 years ago" |
|  | 8 | Married | No information | "Marital Status: Not on file"<br>"Lives with wife, primary support is wife" |
|  | 9 | Married | No information | "Marital status: Married Spouse name: [redacted]" |
| Alcohol Use | 10 | Current User | Current User | "Alcohol use: Yes. Alcohol/week: 6.0 standard drinks. Types: 6 Cans of beer per week. Comment: 3 beers a day on weekend." |
|  | 11 | Never User | Past User | "Denies Tobacco and alcohol use" |
|  | 12 | Current User | Never User | "Alcohol use: Never Alcohol/week: 1.0 standard drink Types: 1 Glasses of wine per week" |
| Tobacco Use | 13 | Past User | Past User | "Smoking status: Former Smoker, Quit date: [year redacted], Years since quitting: 54.4" |
|  | 14 | Past User | No information | "Tobacco Use • Smoking status: Former Smoker Packs/day: 1.50 Years: 32.00 Pack years: 48.00 Types: Cigarettes Quit date: [redacted] Years since quitting: >10 yrs" |
| Drug Use | 15 | Never User | Never User | "He does not use alcohol or any illicit drugs" |
|  | 16 | Current User | Past User | "Drug use: Yes Types: Marijuana Comment: Occasionally smokes a joint (several years ago)" |

\* Only segments within a note that contain effective information about the shown category are shown as examples.
\*\* Company name was masked for publication purpose. SBDH-Reader was fed with real company name when processing.

**Supplementary Table 6.** Performance of SBDH-Reader on development and validation datasets after exclusion of records assigned the “no information” attribute class for each SBDH category. Note-level results are presented as median followed by (lower–upper) bounds of the 95% confidence interval, estimated by bootstrapping with 500 iterations, sampling the entire dataset with replacement.

|  | Macro average |  |  | Adverse attributes only |  |  |
| --- | --- | --- | --- | --- | --- | --- |
| Category | F1 | Precision | Recall | F1 | Precision | Recall |
| MIMIC-G |  |  |  |  |  |  |
| Employment | 1.00<br>(1.00–1.00) | 1.00<br>(1.00–1.00) | 1.00<br>(1.00–1.00) | 1.00<br>(1.00–1.00) | 1.00<br>(1.00–1.00) | 1.00<br>(1.00–1.00) |
| Marital Relationship | 0.92<br>(0.85–0.97) | 0.96<br>(0.88–1.00) | 0.89<br>(0.80–0.96) | 0.94<br>(0.83–1.00) | 0.94<br>(0.80–1.00) | 0.94<br>(0.79–1.00) |
| Overall* | 0.96<br>(0.92–0.99) | 0.98<br>(0.94–1.00) | 0.94<br>(0.90–0.98) | 0.97<br>(0.91–1.00) | 0.97<br>(0.89–1.00) | 0.97<br>(0.89–1.00) |
| MIMIC-A |  |  |  |  |  |  |
| Employment | 0.90<br>(0.89–0.91) | 0.95<br>(0.95–0.96) | 0.86<br>(0.84–0.87) | 0.91<br>(0.90–0.92) | 0.94<br>(0.93–0.95) | 0.88<br>(0.86–0.89) |
| Housing | 0.94<br>(0.91–0.97) | 0.93<br>(0.88–0.96) | 0.96<br>(0.93–0.98) | 0.89<br>(0.84–0.94) | 0.85<br>(0.77–0.91) | 0.94<br>(0.89–0.98) |
| Alcohol Use | 0.98<br>(0.97–0.98) | 0.98<br>(0.98–0.99) | 0.97<br>(0.97–0.98) | 0.98<br>(0.97–0.98) | 0.98<br>(0.97–0.98) | 0.98<br>(0.97–0.98) |
| Tobacco Use | 0.99<br>(0.98–0.99) | 0.99<br>(0.99–0.99) | 0.98<br>(0.98–0.99) | 0.99<br>(0.99–0.99) | 0.99<br>(0.99–0.99) | 0.99<br>(0.98–0.99) |
| Drug Use | 0.94<br>(0.92–0.95) | 0.95<br>(0.94–0.96) | 0.92<br>(0.91–0.94) | 0.90<br>(0.87–0.92) | 0.92<br>(0.89–0.95) | 0.87<br>(0.84–0.90) |
| Overall* | 0.95<br>(0.94–0.95) | 0.96<br>(0.95–0.97) | 0.94<br>(0.93–0.95) | 0.93<br>(0.92–0.94) | 0.94<br>(0.92–0.95) | 0.93<br>(0.92–0.94) |
| UTSW |  |  |  |  |  |  |
| Employment | 0.93<br>(0.90–0.96) | 0.98<br>(0.97–0.99) | 0.89<br>(0.85–0.93) | 0.97<br>(0.95–0.98) | 0.96<br>(0.93–0.98) | 0.98<br>(0.96–0.99) |
| Housing | 0.98<br>(0.95–1.00) | 1.00<br>(0.99–1.00) | 0.96<br>(0.91–0.99) | 0.98<br>(0.92–1.00) | 1.00<br>(1.00–1.00) | 0.96<br>(0.85–1.00) |
| Marital Relationship | 0.98<br>(0.97–0.99) | 0.99<br>(0.98–0.99) | 0.97<br>(0.96–0.98) | 0.98<br>(0.98–0.99) | 0.97<br>(0.96–0.99) | 0.99<br>(0.99–1.00) |
| Alcohol Use | 0.98<br>(0.97–0.99) | 0.98<br>(0.96–0.99) | 0.98<br>(0.97–0.99) | 0.98<br>(0.98–0.99) | 1.00<br>(0.99–1.00) | 0.97<br>(0.95–0.98) |
| Tobacco Use | 0.99<br>(0.99–1.00) | 1.00<br>(0.99–1.00) | 0.99<br>(0.99–1.00) | 0.99<br>(0.99–1.00) | 1.00<br>(1.00–1.00) | 0.99<br>(0.98–1.00) |
| Drug Use | 0.98<br>(0.97–0.99) | 0.99<br>(0.99–1.00) | 0.97<br>(0.95–0.98) | 0.97<br>(0.95–0.98) | 1.00<br>(1.00–1.00) | 0.94<br>(0.91–0.97) |
| Overall | 0.97<br>(0.96–0.98) | 0.99<br>(0.98–0.99) | 0.96<br>(0.96–0.97) | 0.98<br>(0.97–0.99) | 0.99<br>(0.98–0.99) | 0.97<br>(0.95–0.98) |

\* The overall metrics report macro-average across all SBDH categories.

**Supplementary Table 7.**
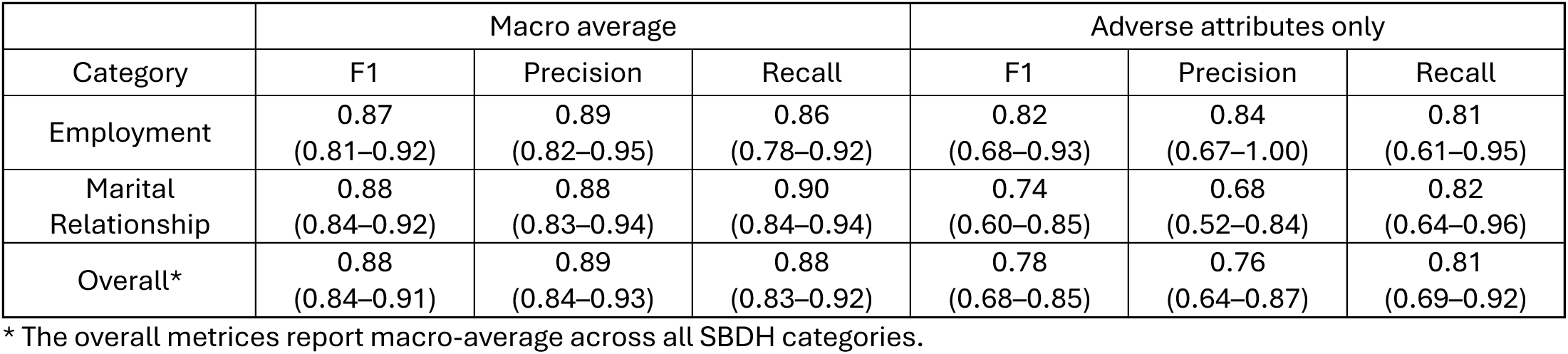
Performance of SBDH-Reader on sentence-level inputs from the MIMIC-G dataset. Sentence-level results are presented as median followed by (lower–upper) bounds of the 95% confidence interval, estimated by bootstrapping with 500 iterations, sampling the entire dataset with replacement.

## References

1. Marmot M, Friel S, Bell R, Houweling TA, Taylor S, Commission on Social Determinants of Health. Closing the gap in a generation: health equity through action on the social determinants of health. Lancet. Nov 8 2008;372(9650):1661–9. doi:10.1016/S0140-6736(08)61690-6

2. Adler NE, Glymour MM, Fielding J. Addressing Social Determinants of Health and Health Inequalities. JAMA. Oct 25 2016;316(16):1641–1642. doi:10.1001/jama.2016.14058

3. Davis R, Campbell R, Hildon Z, Hobbs L, Michie S. Theories of behaviour and behaviour change across the social and behavioural sciences: a scoping review. Health Psychol Rev. 2015;9(3):323–44. doi:10.1080/17437199.2014.941722

4. Chen M, Tan X, Padman R. Social determinants of health in electronic health records and their impact on analysis and risk prediction: A systematic review. J Am Med Inform Assoc. Nov 1 2020;27(11):1764–1773. doi:10.1093/jamia/ocaa143

5. Lu MLR, Davila CD, Shah M, et al. Marital status and living condition as predictors of mortality and readmissions among African Americans with heart failure. International Journal of Cardiology. Nov 1 2016;222:313–318. doi:10.1016/j.ijcard.2016.07.185

6. Sterling MR, Ringel JB, Pinheiro LC, et al. Social Determinants of Health and 90-Day Mortality After Hospitalization for Heart Failure in the REGARDS Study. J Am Heart Assoc. May 5 2020;9(9):e014836. doi:10.1161/JAHA.119.014836

7. Segar MW, Hall JL, Jhund PS, et al. Machine Learning-Based Models Incorporating Social Determinants of Health vs Traditional Models for Predicting In-Hospital Mortality in Patients With Heart Failure. JAMA Cardiol. Aug 1 2022;7(8):844–854. doi:10.1001/jamacardio.2022.1900

8. Hiatt RA, Breen N. The social determinants of cancer - A challenge for transdisciplinary science. American Journal of Preventive Medicine. Aug 2008;35(2):S141–S150. doi:10.1016/j.amepre.2008.05.006

9. Adkins-Jackson PB, George KM, Besser LM, et al. The structural and social determinants of Alzheimer’s disease related dementias. Alzheimers Dement. Jul 2023;19(7):3171–3185. doi:10.1002/alz.13027

10. Sekar RR, Herrel LA, Stensland KD. Social Determinants of Health and the Availability of Cancer Clinical Trials in the United States. JAMA Netw Open. May 1 2024;7(5):e2410162. doi:10.1001/jamanetworkopen.2024.10162

11. Rae S, Shaya S, Taylor E, et al. Social determinants of health inequalities in early phase clinical trials in Northern England. Br J Cancer. Sep 2024;131(4):685–691. doi:10.1038/s41416-024-02765-w

12. Hatef E, Rouhizadeh M, Tia I, et al. Assessing the Availability of Data on Social and Behavioral Determinants in Structured and Unstructured Electronic Health Records: A Retrospective Analysis of a Multilevel Health Care System. JMIR Med Inform. Aug 2 2019;7(3):e13802. doi:10.2196/13802

13. Truong HP, Luke AA, Hammond G, Wadhera RK, Reidhead M, Joynt Maddox KE. Utilization of Social Determinants of Health ICD-10 Z-Codes Among Hospitalized Patients in the United States, 2016-2017. Med Care. Dec 2020;58(12):1037–1043. doi:10.1097/MLR.0000000000001418

14. Dupre ME, Nelson A, Lynch SM, et al. Socioeconomic, Psychosocial and Behavioral Characteristics of Patients Hospitalized With Cardiovascular Disease. Am J Med Sci. Dec 2017;354(6):565–572. doi:10.1016/j.amjms.2017.07.011

15. Dupre ME, Nelson A, Lynch SM, et al. Identifying Nonclinical Factors Associated With 30-Day Readmission in Patients with Cardiovascular Disease: Protocol for an Observational Study. JMIR Res Protoc. Jun 15 2017;6(6):e118. doi:10.2196/resprot.7434

16. Guo Y, Chen Z, Xu K, et al. International Classification of Diseases, Tenth Revision, Clinical Modification social determinants of health codes are poorly used in electronic health records. Medicine (Baltimore). Dec 24 2020;99(52):e23818. doi:10.1097/MD.0000000000023818

17. Vest JR, Grannis SJ, Haut DP, Halverson PK, Menachemi N. Using structured and unstructured data to identify patients’ need for services that address the social determinants of health. Int J Med Inform. Nov 2017;107:101–106. doi:10.1016/j.ijmedinf.2017.09.008

18. Patra BG, Sharma MM, Vekaria V, et al. Extracting social determinants of health from electronic health records using natural language processing: a systematic review. J Am Med Inform Assoc. Nov 25 2021;28(12):2716–2727. doi:10.1093/jamia/ocab170

19. Mullangi S, Aviki EM, Hershman DL. Reexamining Social Determinants of Health Data Collection in the COVID-19 Era. JAMA Oncol. Dec 1 2022;8(12):1736–1738. doi:10.1001/jamaoncol.2022.4543

20. Ong JCL, Seng BJJ, Law JZF, et al. Artificial intelligence, ChatGPT, and other large language models for social determinants of health: Current state and future directions. Cell Rep Med. Jan 16 2024;5(1):101356. doi:10.1016/j.xcrm.2023.101356

21. Bompelli A, Wang Y, Wan R, et al. Social and Behavioral Determinants of Health in the Era of Artificial Intelligence with Electronic Health Records: A Scoping Review. Health Data Sci. 2021;2021:9759016. doi:10.34133/2021/9759016

22. Dorr D, Bejan CA, Pizzimenti C, Singh S, Storer M, Quinones A. Identifying Patients with Significant Problems Related to Social Determinants of Health with Natural Language Processing. Stud Health Technol Inform. Aug 21 2019;264:1456–1457. doi:10.3233/SHTI190482

23. Reeves RM, Christensen L, Brown JR, et al. Adaptation of an NLP system to a new healthcare environment to identify social determinants of health. J Biomed Inform. Aug 2021;120:103851. doi:10.1016/j.jbi.2021.103851

24. Lybarger K, Ostendorf M, Yetisgen M. Annotating social determinants of health using active learning, and characterizing determinants using neural event extraction. J Biomed Inform. Jan 2021;113:103631. doi:10.1016/j.jbi.2020.103631

25. Han S, Zhang RF, Shi L, et al. Classifying social determinants of health from unstructured electronic health records using deep learning-based natural language processing. J Biomed Inform. Mar 2022;127:103984. doi:10.1016/j.jbi.2021.103984

26. Raza S, Dolatabadi E, Ondrusek N, Rosella L, Schwartz B. Discovering social determinants of health from case reports using natural language processing: algorithmic development and validation. BMC Digital Health. 2023/09/11 2023;1(1):35. doi:10.1186/s44247-023-00035-y

27. Wu W, Holkeboer KJ, Kolawole TO, Carbone L, Mahmoudi E. Natural language processing to identify social determinants of health in Alzheimer’s disease and related dementia from electronic health records. Health Serv Res. Dec 2023;58(6):1292–1302. doi:10.1111/1475-6773.14210

28. Roy S, Morrell S, Zhao L, Homayouni R. Large-scale identification of social and behavioral determinants of health from clinical notes: comparison of Latent Semantic Indexing and Generative Pretrained Transformer (GPT) models. BMC Med Inform Decis Mak. Oct 10 2024;24(1):296. doi:10.1186/s12911-024-02705-x

29. Guevara M, Chen S, Thomas S, et al. Large language models to identify social determinants of health in electronic health records. NPJ Digit Med. Jan 11 2024;7(1):6. doi:10.1038/s41746-023-00970-0

30. Gabriel RA, Litake O, Simpson S, Burton BN, Waterman RS, Macias AA. On the development and validation of large language model-based classifiers for identifying social determinants of health. Proc Natl Acad Sci U S A. Sep 24 2024;121(39):e2320716121. doi:10.1073/pnas.2320716121

31. Keloth VK, Selek S, Chen Q, et al. Large Language Models for Social Determinants of Health Information Extraction from Clinical Notes - A Generalizable Approach across Institutions. medRxiv [Preprint]. May 22 2024;doi:10.1101/2024.05.21.24307726

32. Lybarger K, Yetisgen M, Uzuner O. The 2022 n2c2/UW shared task on extracting social determinants of health. J Am Med Inform Assoc. Jul 19 2023;30(8):1367–1378. doi:10.1093/jamia/ocad012

33. Feller DJ, Bear Don’t Walk Iv OJ, Zucker J, Yin MT, Gordon P, Elhadad N. Detecting Social and Behavioral Determinants of Health with Structured and Free-Text Clinical Data. Appl Clin Inform. Jan 2020;11(1):172–181. doi:10.1055/s-0040-1702214

34. Thirunavukarasu AJ, Ting DSJ, Elangovan K, Gutierrez L, Tan TF, Ting DSW. Large language models in medicine. Nat Med. Aug 2023;29(8):1930–1940. doi:10.1038/s41591-023-02448-8

35. Will ChatGPT transform healthcare? Nat Med. Mar 2023;29(3):505–506. doi:10.1038/s41591-023-02289-5

36. Patel SB, Lam K. ChatGPT: the future of discharge summaries? Lancet Digit Health. Mar 2023;5(3):e107–e108. doi:10.1016/S2589-7500(23)00021-3

37. Shah NH, Entwistle D, Pfeffer MA. Creation and Adoption of Large Language Models in Medicine. JAMA. Sep 5 2023;330(9):866–869. doi:10.1001/jama.2023.14217

38. Ayers JW, Poliak A, Dredze M, et al. Comparing Physician and Artificial Intelligence Chatbot Responses to Patient Questions Posted to a Public Social Media Forum. JAMA Intern Med. Jun 1 2023;183(6):589–596. doi:10.1001/jamainternmed.2023.1838

39. Harrer S. Attention is not all you need: the complicated case of ethically using large language models in healthcare and medicine. EBioMedicine. Apr 2023;90:104512. doi:10.1016/j.ebiom.2023.104512

40. Yang X, Chen A, PourNejatian N, et al. A large language model for electronic health records. NPJ Digit Med. Dec 26 2022;5(1):194. doi:10.1038/s41746-022-00742-2

41. Huang J, Yang DM, Rong R, et al. A critical assessment of using ChatGPT for extracting structured data from clinical notes. NPJ Digit Med. May 1 2024;7(1):106. doi:10.1038/s41746-024-01079-8

42. Nezafati K, Wang L, Rong R, et al. Assessing disease severity in cutaneous lupus patients using natural language processing: Preliminary data from a cohort study. J Am Acad Dermatol. Nov 28 2024;doi:10.1016/j.jaad.2024.10.105

43. Stemerman R, Arguello J, Brice J, Krishnamurthy A, Houston M, Kitzmiller R. Identification of social determinants of health using multi-label classification of electronic health record clinical notes. JAMIA Open. Jul 2021;4(3):ooaa069. doi:10.1093/jamiaopen/ooaa069

44. Google. Prompt engineering: overview and guide. Accessed 5/21/2025, 2025. https://cloud.google.com/discover/what-is-prompt-engineering#prompt-engineering-overview-and-guide

45. Yu Z, Peng C, Yang X, et al. Identifying social determinants of health from clinical narratives: A study of performance, documentation ratio, and potential bias. J Biomed Inform. May 2024;153:104642. doi:10.1016/j.jbi.2024.104642

46. Johnson AE, Pollard TJ, Shen L, et al. MIMIC-III, a freely accessible critical care database. Sci Data. May 24 2016;3:160035. doi:10.1038/sdata.2016.35

47. Ahsan H, Ohnuki E, Mitra A, Yu H. MIMIC-SBDH: A Dataset for Social and Behavioral Determinants of Health. Proc Mach Learn Res. Aug 2021;149:391–413.

48. PhysioNet. PhysioNet Credentialed Health Data License 1.5.0. Accessed 2/19/2025, https://physionet.org/about/licenses/physionet-credentialed-health-data-license-150/

49. PhysioNet. Responsible use of MIMIC data with online services like GPT. Accessed 1/19/2025, 2025. https://physionet.org/news/post/gpt-responsible-use

50. Klie JC, Bugert M, Boullosa B, Eckart de Castilho R, Gurevych I. The INCEpTION Platform: Machine-Assisted and Knowledge-Oriented Interactive Annotation. presented at: In Proceedings of System Demonstrations of the 27th International Conference on Computational Linguistics; 2018; Santa Fe, NM.

